# HLA Class II Alleles are associated with Anti-Drug Antibodies in Ulcerative Colitis Patients Treated with Etrolizumab

**DOI:** 10.1101/2025.03.28.25320486

**Authors:** Ashis Saha, Mark I. McCarthy, Saloumeh K. Fischer, Gizette Sperinde, Christian Hammer

## Abstract

Patients treated with etrolizumab, a monoclonal antibody drug for ulcerative colitis, often develop anti-drug antibodies (ADA) and neutralizing antibodies (NAb). To investigate genetic risk factors, we evaluated associations between HLA alleles and ADA/NAb development. *HLA-DQB1*06:03* demonstrated the most significant association with both ADA (OR = 7.3, P ≤ 7.6×10^−13^) and NAb (OR = 13.1, P ≤ 7.7×10^−13^). After controlling for *HLA-DQB1*06:03, HLA-DQA1*03:03* emerged as the next most significant allele associated with ADA (OR = 2.8, P ≤ 1.0×10^−3^) and NAb (OR = 5.0, P ≤ 1.3×10^−3^). Notably, all patients carrying both alleles (n=5) developed ADA, suggesting these two alleles are sufficient to induce ADA to etrolizumab.

## Introduction

Etrolizumab, a monoclonal antibody targeting the β7 subunit of α4β7 and αEβ7 integrin heterodimers, was developed for the treatment of ulcerative colitis^1–3^ and evaluated in Phase III clinical trials. However, these trials were discontinued due to mixed efficacy results. A fraction of patients treated with etrolizumab developed undesirable immune responses, characterized by the formation of anti-drug antibodies (ADA), a subset of which were neutralizing antibodies (NAb). ADAs can alter pharmacokinetics and induce adverse reactions, thereby impacting the overall therapeutic efficacy and safety profiles. NAbs can neutralize the therapeutic effects of the drug, further complicating the therapeutic landscape.

Inherited genetic variation in Human Leukocyte Antigen (HLA) molecules is known to play a role in ADA development, given their function in presenting peptide antigens to T cells. Previous studies reported associations between specific HLA class II alleles and the risk of ADA development during treatment with therapeutic proteins such as interferon beta (IFNβ), anti-PD-L1, and anti-tumor necrosis factor (anti-TNF) antibodies^4–6^. For example, the *HLA-DQA1*05* allele was associated with ADA development (hazard ratio (HR) = 1.9, P ≤ 5.9×10^−13^) in Crohn’s disease patients treated with infliximab or adalimumab^5^. Similarly, *HLA-DRB1*01:01* was associated with ADA (odds ratio (OR) = 2.0, P ≤ 3.4×10^−5^) and NAb development (OR = 2.3, P ≤ 2.8×10^−7^) in cancer patients treated with atezolizumab^6^.

Based on evidence from monoclonal antibody treatments such as infliximab, adalimumab, and atezolizumab, we investigated whether the variable presentation of etrolizumab peptides via HLA molecules contributes to ADA formation. Here, we present a genetic association study involving 615 patients treated with etrolizumab for ulcerative colitis across three different phase III clinical trials: Hickory^1^, Laurel^2^, and Gardenia^3^. The impact of etrolizumab-associated ADAs and NAbs development on clinical outcomes is currently under investigation and will be reported separately. In this study, we focus on understanding the baseline genetic characteristics of patients that may predispose them to developing ADAs or NAbs.

## Materials and methods

### Studies and subjects

This research utilized samples from participants who consented to genetic analyses in Roche sponsored clinical trials: Hickory (NCT02100696), Laurel (NCT02165215), and Gardenia (NCT02136069). In total we included 615 patients of European ancestry across these three studies.

### HLA typing

For the patients in the Hickory and Laurel clinical trials, genomic DNA was extracted from blood samples using the DNA Blood400 kit (Chemagic) and eluted in 50μL Elution Buffer (EB, Qiagen). DNA was sheared (Covaris LE220) and sequencing libraries were prepared using the TruSeq Nano DNA HT kit (Illumina Inc.). 150bp paired-end whole-genome sequencing (WGS) data was generated to an average read depth of 30× using the HiSeq platform (Illumina X10, San Diego, CA, USA) and processed using the Burrows Wheeler Aligner (BWA)/Genome Analysis Toolkit (GATK) best practices pipeline. Short reads were mapped to hg38/GRCh38 (GCA_000001405.15), including alternate assemblies, using an alt-aware version of BWA to generate BAM files. All sequencing data was checked for concordance with SNP fingerprint data collected before sequencing. HLA-HD^7^ was used to infer HLA genotypes starting from BAM files generated as described above. HLA calls at 6-digit resolution were reduced to 4-digit level, reflecting the amino-acid sequence of the HLA protein.

For the Gardenia study, samples were genotyped with the Infinium Global Screen Array (v2) by Illumina and imputed using BEAGLE (v5.1)^8^ and the 1000 Genomes reference panel (GRCh38 aligned version). HLA alleles were imputed using the HIBAG^9^ software with ancestry-specific reference panels.

### ADA and NAb bioanalytical assays

Anti-etrolizumab antibodies were detected and characterized using a tiered strategy. Initially, serum samples were screened using a validated bridging enzyme-linked immunosorbent assay (ELISA) assay. Positive samples were then confirmed through competitive binding with etrolizumab (10 µg/mL). Once confirmed, these samples were further characterized for their neutralization activity using the neutralizing ADA (NAb) assay.

### Statistical analyses

For each clinical trial, we performed logistic regression to assess the association between ADA or NAb and HLA alleles with carrier frequencies between 2% and 98%, assuming a dominant inheritance model. We included age, sex, and three principal components (PCs) as covariates to adjust for population stratification. We conducted a random-effects meta-analysis using the R package “meta” to calculate effect estimates, 95% confidence intervals, and p-values. We applied the Benjamini-Hochberg method to correct for multiple testing.

## Results

ADA measures for each patient were obtained using a bridging enzyme-linked immunosorbent assay (ELISA) with positives confirmed via competitive binding with etrolizumab. Neutralization activities of confirmed positive samples were characterized with a NAb assay. Among 615 patients of European ancestry who provided consent for genetic analyses, 182 (29.6%) patients developed treatment emergent anti-drug antibodies (ADAs) to etrolizumab across the three studies (Table 1). Seventy-seven of the 182 ADA positive patients also developed neutralizing antibodies (see supplementary materials for details).

**Table 1.**
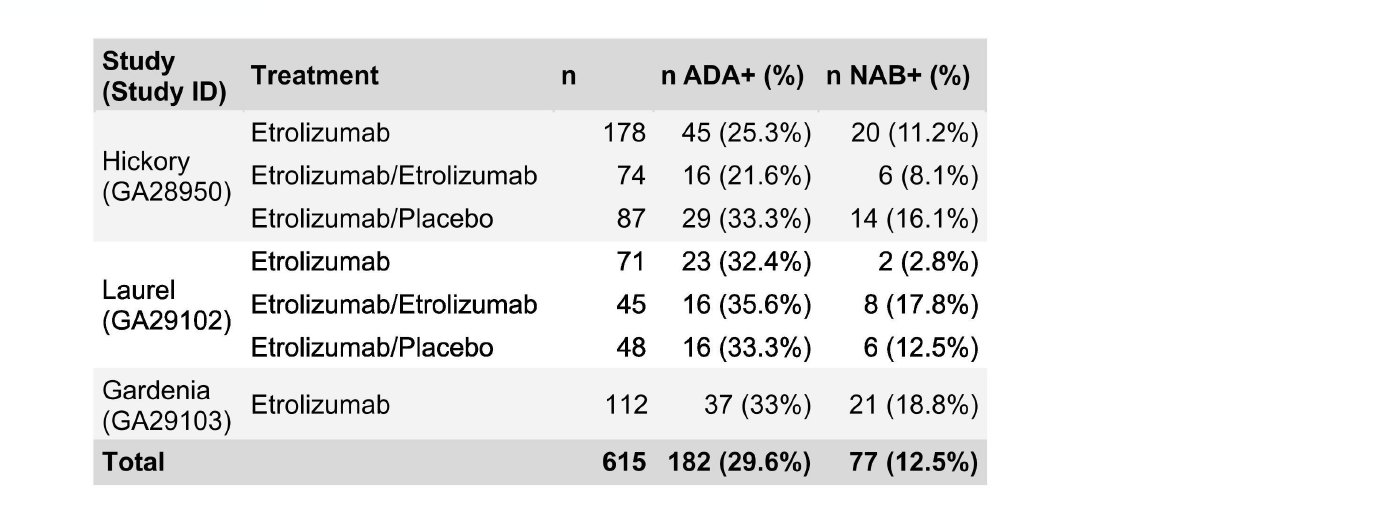
Number of patients (n), ADA and NAb rates according to study and treatment arm. Patients in Etrolizumab arms were treated with etrolizumab. Patients in Etrolizumab/Etrolizumab arms were treated with Etrolizumab in both induction and maintenance phase. Patients in Etrolizumab/Placebo arms were treated with Etrolizumab in the induction phase, but not in the maintenance phase.

| Study<br>(Study ID) | Treatment | n | n ADA+ (%) | n NAB+ (%) |
| --- | --- | --- | --- | --- |
| Hickory<br>(GA28950) | Etrolizumab | 178 | 45 (25.3%) | 20 (11.2%) |
|  | Etrolizumab/Etrolizumab | 74 | 16 (21.6%) | 6 (8.1%) |
|  | Etrolizumab/Placebo | 87 | 29 (33.3%) | 14 (16.1%) |
| Laurel<br>(GA29102) | Etrolizumab | 71 | 23 (32.4%) | 2 (2.8%) |
|  | Etrolizumab/Etrolizumab | 45 | 16 (35.6%) | 8 (17.8%) |
|  | Etrolizumab/Placebo | 48 | 16 (33.3%) | 6 (12.5%) |
| Gardenia<br>(GA29103) | Etrolizumab | 112 | 37 (33%) | 21 (18.8%) |
| **Total** |  | **615** | **182 (29.6%)** | **77 (12.5%)** |

We applied logistic regression on each study to test for association between ADA status and HLA alleles (4-digit resolution). We limited analyses to HLA alleles with carrier frequencies between 2% and 98% (minimizing the multiple testing burden that would result from adding lots of alleles with limited power). Age, sex, and three genetic principal components were included as covariates. A meta-analysis, using a random-effects model implemented in the R package “meta”, identified statistically significant associations (FDR ≤ 0.05 after Benjamini-Hochberg correction for multiple testing) between multiple HLA class II alleles and ADA occurrence (Figure 1, Supplemental Table 1). Of these alleles, *HLA-DQB1*06:03* demonstrated the most significant association (OR = 7.3, 95% CI = [4.2, 12.6], P ≤ 7.6×10^−13^, FDR ≤ 5.0×10^−11^), with each clinical trial showing an odds ratio greater than 6. *HLA-DQB1\**06:03 is part of a common haplotype that encompasses the three top-associated alleles (*HLA-DQB1\**06:03, *HLA-DRB1*13:01, HLA-DQA1*01:03*, Supplemental Table 1), which are therefore not statistically independent. A conditional analysis, including *HLA-DQB1*06:03* as an additional covariate, showed no residual signal at *HLA-DRB1*13:01* and *HLA-DQA1*01:03, and* uncovered an independent association of a different HLA class II allele, *HLA-DQA1*03:03* (OR = 2.8, 95% CI = [1.5, 5.3], P ≤ 1.0×10^−3^, FDR ≤ 0.04, Figure 1, Supplemental Table 2).

**Figure 1.**
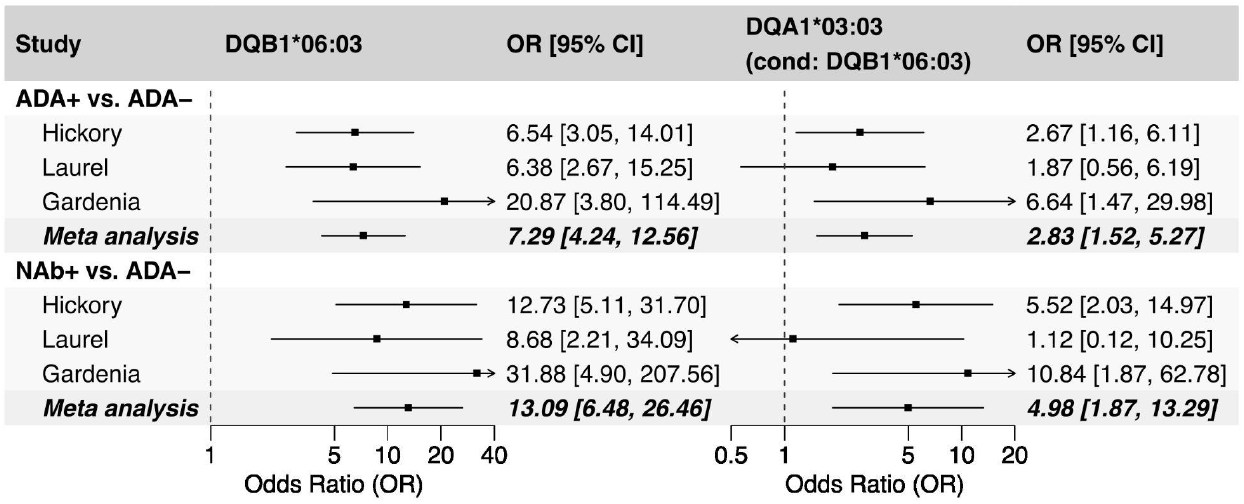
Forest plot showing the effect estimates of the two independent HLA alleles: HLA-DQB1*06:03 (column 2-3) and HLA-DQA1*03:03 (column 4-5). The effects of HLA-DQA1*03:03 were estimated after conditioning on the first independent allele HLA-DQB1*06:03. Bars represent 95% confidence intervals (CI) for odds ratios.

We then conducted a subgroup analysis on patients who developed NAbs, which directly inhibit the biological activity of the drug by binding to its target binding domain. We compared NAb-positive patients to ADA-negative patients, excluding ADA-positive but NAb-negative patients. The analysis revealed that both independent alleles identified in the previous analysis were also significant in this subgroup: *HLA-DQB1*06:03* (OR = 13.1, 95% CI = [6.5, 26.5], P ≤ 7.7×10^−13^, FDR ≤ 4.9×10^−11^) and *HLA-DQA1*03:03* (OR = 5.0, 95% CI = [1.9, 13.3], P ≤ 1.3×10^−3^, FDR ≤ 0.04, Figure 1). This finding suggests that individuals with these alleles have a predisposition to developing immune responses against etrolizumab, affecting both the binding and neutralizing capacities of the drug.

Notably, the risk of developing ADA or NAb increases with the number of unique independent alleles carried by each individual in each study (Figure 2). In particular, all 5 patients carrying both independently associated HLA alleles (*HLA-DQB1*06:03* and *HLA-DQA1*03:03*) were positive for ADA, with 3 of them also positive for Nab. Though numbers are small, these findings may indicate that presence of both alleles is sufficient to induce ADA formation.

**Figure 2.**
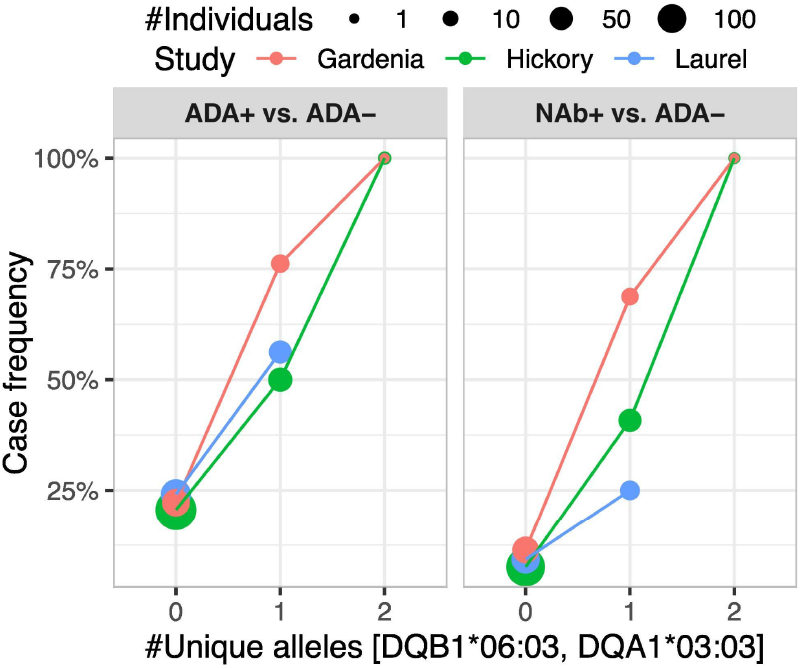
The risk of developing ADA or NAb (y-axis) increases with the number of unique independent alleles carried by each individual (x-axis). A value of 0, 1 and 2 on the x-axis represent individuals carrying none, either and both of the two independent alleles, respectively. The size of the point represents the number of allele carriers.

## Discussion

In the context of a limited understanding of patient factors driving anti-drug antibody (ADA) risk, our study has identified two HLA class II alleles (*HLA-DQB1*06:03* and *HLA-DQA1*03:03*) associated with an increased risk of ADA formation against etrolizumab. Our findings unveil one of the strongest reported associations to date between an HLA allele (*HLA-DQB1*06:03*) and both ADA (P ≤ 7.6×10^−13^) and NAb (P ≤ 7.7×10^−13^) status for antibody therapeutics. Similarly, Sazonovs et al.^5^ reported a strong association (P ≤ 5.9×10^−13^) between *HLA-DQA1*05* and ADA development in Crohn’s disease patients with infliximab or adalimumab therapy. These findings suggest that while HLA plays a crucial role in ADA development, different alleles are involved in distinct patient populations. Further research on the generalizability and specificity of genetic signals across different antibody therapies could provide valuable insights for advancing drug development and enhancing personalized treatment strategies.

The carrier frequencies of the two alleles reported here are close to 10% in people of European ancestry (13.3% for *HLA-DQB1*06:03* and 8.9% for *HLA-DQA1*03:03* in our dataset), which suggests that a considerable subset of the patient population is predisposed to ADA formation to etrolizumab. This prevalence underscores the importance of identifying and potentially eliminating immunogenic epitopes within recombinant large molecule therapeutics.

In addition, two other alleles, *HLA-DRB1*13:01* and *HLA-DQA1*01:03*, which are in linkage disequilibrium with *HLA-DQB1*06:03*, expectedly showed strong associations with ADA development (OR: 6.2 and 3.7, respectively; P ≤ 2.9×10^−9^ and 1.4×10^−7^, respectively). The haplotype containing these alleles likely contributes to ADA risk; however, further studies are required to identify the causal allele(s).

It is important to recognize that HLA alleles only partially explain the variance in ADA formation. Consequently, future research should continue to explore other potential risk factors and molecular mechanisms to better understand and mitigate the impact of ADA on therapeutic efficacy and patient outcomes, potentially leading to more personalized and effective treatment strategies.

## Acknowledgement

Artificial intelligence (GPT-4) was used to edit texts.

## Ethics statement

The research was performed with samples from subjects who had given consent for genetic research in Roche sponsored clinical trials: Hickory (NCT02100696), Laurel (NCT02165215), and Gardenia (NCT02136069). In each of these trials, the Ethics Committees (EC) and Institutional Review Boards (IRB) who approved the trials also approved the Informed Consent Forms (ICF) used to obtain consent for genetic research from the study participants. No EC/IRB was additionally consulted to approve the specific genetic research reported here but an internal team of consent experts made sure the genetic research is covered by the ICFs signed by the study participants. Complete lists of ECs and IRBs are available in the supplementary materials (Supplemental Table 3-5).

## Data availability statement

HLA association summary statistics are available in the supplementary materials. Qualified researchers may request access to individual patient data used in this study through Roche data sharing platforms in accordance with the Global Policy on Sharing of Clinical Study Information: http://www.roche.com/research_and_development/who_we_are_how_we_work/clinical_trials/our_commitment_to_data_sharing.htm.

