## Supplementary materials for "HLA Class II Alleles are associated with Anti-Drug Antibodies in Ulcerative Colitis Patients Treated with Etrolizumab"

***Supplemental Table 1: Meta-analysis results***

| Analysis | allele | OR | conf.lower | conf.upper | seTE | pval | pval.bh |
|---|---|---|---|---|---|---|---|
| ADA+ vs. ADA- | DQB1*06:03 | 7.2935 | 4.2362 | 12.5575 | 1.3194 | 7.63E-13 | 5.03E-11 |
| ADA+ vs. ADA- | DRB1*13:01 | 6.2116 | 3.3988 | 11.3523 | 1.3602 | 2.91E-09 | 9.61E-08 |
| ADA+ vs. ADA- | DQA1*01:03 | 3.6831 | 2.2668 | 5.9843 | 1.2810 | 1.41E-07 | 3.09E-06 |
| ADA+ vs. ADA- | DRB1*03:01 | 0.3774 | 0.2100 | 0.6782 | 1.3486 | 1.12E-03 | 1.85E-02 |
| ADA+ vs. ADA- | DQA1*05:01 | 0.3992 | 0.2248 | 0.7092 | 1.3406 | 1.73E-03 | 2.06E-02 |
| ADA+ vs. ADA- | B*15:01 | 2.3584 | 1.3733 | 4.0499 | 1.3177 | 1.87E-03 | 2.06E-02 |
| ADA+ vs. ADA- | DQB1*02:01 | 0.3813 | 0.2041 | 0.7122 | 1.3755 | 2.49E-03 | 2.35E-02 |
| ADA+ vs. ADA- | A*26:01 | 2.5985 | 1.3230 | 5.1037 | 1.4112 | 5.56E-03 | 4.59E-02 |
| ADA+ vs. ADA- | A*68:01 | 2.3900 | 1.1864 | 4.8147 | 1.4295 | 1.48E-02 | 1.08E-01 |
| ADA+ vs. ADA- | A*01:01 | 0.5970 | 0.3878 | 0.9191 | 1.2462 | 1.91E-02 | 1.26E-01 |
| ADA+ vs. ADA- | DQA1*03:03 | 2.4092 | 1.1052 | 5.2517 | 1.4882 | 2.70E-02 | 1.62E-01 |
| ADA+ vs. ADA- | C*03:04 | 1.8777 | 1.0578 | 3.3330 | 1.3401 | 3.14E-02 | 1.73E-01 |
| ADA+ vs. ADA- | DQB1*05:01 | 0.6185 | 0.3938 | 0.9715 | 1.2591 | 3.70E-02 | 1.88E-01 |
| ADA+ vs. ADA- | DQA1*01:01 | 0.6215 | 0.3908 | 0.9884 | 1.2671 | 4.45E-02 | 1.97E-01 |
| ADA+ vs. ADA- | B*08:01 | 0.4366 | 0.1939 | 0.9830 | 1.5129 | 4.54E-02 | 1.97E-01 |
| ADA+ vs. ADA- | C*03:03 | 1.9236 | 1.0063 | 3.6772 | 1.3918 | 4.78E-02 | 1.97E-01 |
| ADA+ vs. ADA- | C*07:01 | 0.6602 | 0.4345 | 1.0031 | 1.2379 | 5.17E-02 | 2.01E-01 |
| ADA+ vs. ADA- | DRB1*04:01 | 2.5308 | 0.9313 | 6.8777 | 1.6654 | 6.87E-02 | 2.52E-01 |
| ADA+ vs. ADA- | DQA1*05:05 | 0.5953 | 0.3173 | 1.1170 | 1.3786 | 1.06E-01 | 3.69E-01 |
| ADA+ vs. ADA- | DRB1*11:01 | 0.6420 | 0.3690 | 1.1169 | 1.3265 | 1.17E-01 | 3.85E-01 |
| ADA+ vs. ADA- | C*12:03 | 2.2271 | 0.8060 | 6.1534 | 1.6796 | 1.23E-01 | 3.85E-01 |
| ADA+ vs. ADA- | B*52:01 | 0.4862 | 0.1862 | 1.2695 | 1.6319 | 1.41E-01 | 4.00E-01 |
| ADA+ vs. ADA- | DPA1*01:03 | 0.5075 | 0.2026 | 1.2711 | 1.5974 | 1.48E-01 | 4.00E-01 |
| ADA+ vs. ADA- | DPA1*02:01 | 1.3852 | 0.8906 | 2.1546 | 1.2528 | 1.48E-01 | 4.00E-01 |
| ADA+ vs. ADA- | DRA*01:02 | 1.3198 | 0.9033 | 1.9285 | 1.2135 | 1.52E-01 | 4.00E-01 |
| ADA+ vs. ADA- | DQA1*01:02 | 1.2879 | 0.8835 | 1.8773 | 1.2120 | 1.88E-01 | 4.74E-01 |
| ADA+ vs. ADA- | DRB1*11:04 | 0.6481 | 0.3368 | 1.2471 | 1.3964 | 1.94E-01 | 4.74E-01 |
| ADA+ vs. ADA- | A*25:01 | 1.6287 | 0.7565 | 3.5064 | 1.4788 | 2.13E-01 | 4.80E-01 |
| ADA+ vs. ADA- | C*05:01 | 1.4068 | 0.8210 | 2.4107 | 1.3163 | 2.14E-01 | 4.80E-01 |
| ADA+ vs. ADA- | DRB1*15:01 | 1.3961 | 0.8179 | 2.3829 | 1.3136 | 2.21E-01 | 4.80E-01 |

| | | | | | | | |
|---|---|---|---|---|---|---|---|
| ADA+ vs. ADA- | C*08:02 | 0.5841 | 0.2450 | 1.3929 | 1.5580 | 2.25E-01 | 4.80E-01 |
| ADA+ vs. ADA- | B*14:02 | 0.5755 | 0.2257 | 1.4674 | 1.6121 | 2.47E-01 | 5.10E-01 |
| ADA+ vs. ADA- | A*02:01 | 1.2262 | 0.8537 | 1.7611 | 1.2029 | 2.70E-01 | 5.29E-01 |
| ADA+ vs. ADA- | B*40:01 | 1.4448 | 0.7486 | 2.7882 | 1.3986 | 2.73E-01 | 5.29E-01 |
| ADA+ vs. ADA- | B*38:01 | 2.3207 | 0.4851 | 11.1026 | 2.2225 | 2.92E-01 | 5.38E-01 |
| ADA+ vs. ADA- | C*02:02 | 0.6456 | 0.2853 | 1.4609 | 1.5169 | 2.94E-01 | 5.38E-01 |
| ADA+ vs. ADA- | DQB1*05:03 | 0.6998 | 0.3328 | 1.4717 | 1.4612 | 3.47E-01 | 6.09E-01 |
| ADA+ vs. ADA- | B*44:02 | 1.2920 | 0.7546 | 2.2122 | 1.3157 | 3.50E-01 | 6.09E-01 |
| ADA+ vs. ADA- | DQA1*01:04 | 0.7178 | 0.3426 | 1.5040 | 1.4585 | 3.80E-01 | 6.42E-01 |
| ADA+ vs. ADA- | DRB1*01:01 | 0.7918 | 0.4630 | 1.3539 | 1.3148 | 3.94E-01 | 6.49E-01 |
| ADA+ vs. ADA- | A*24:02 | 0.8018 | 0.4770 | 1.3476 | 1.3033 | 4.04E-01 | 6.51E-01 |
| ADA+ vs. ADA- | A*03:01 | 1.2680 | 0.7132 | 2.2544 | 1.3412 | 4.19E-01 | 6.58E-01 |
| ADA+ vs. ADA- | B*18:01 | 1.2620 | 0.6838 | 2.3294 | 1.3671 | 4.57E-01 | 6.96E-01 |
| ADA+ vs. ADA- | DRB1*13:02 | 1.3337 | 0.6157 | 2.8889 | 1.4834 | 4.65E-01 | 6.96E-01 |
| ADA+ vs. ADA- | DQA1*02:01 | 1.1757 | 0.7463 | 1.8523 | 1.2610 | 4.85E-01 | 6.96E-01 |
| ADA+ vs. ADA- | DRB1*07:01 | 1.1757 | 0.7463 | 1.8523 | 1.2610 | 4.85E-01 | 6.96E-01 |
| ADA+ vs. ADA- | B*35:01 | 0.7409 | 0.3070 | 1.7883 | 1.5676 | 5.05E-01 | 7.09E-01 |
| ADA+ vs. ADA- | DRB1*14:54 | 0.7793 | 0.3535 | 1.7177 | 1.4967 | 5.36E-01 | 7.29E-01 |
| ADA+ vs. ADA- | B*51:01 | 1.3142 | 0.5468 | 3.1584 | 1.5642 | 5.41E-01 | 7.29E-01 |
| ADA+ vs. ADA- | A*32:01 | 0.8204 | 0.4202 | 1.6015 | 1.4068 | 5.62E-01 | 7.42E-01 |
| ADA+ vs. ADA- | B*35:03 | 0.7890 | 0.3396 | 1.8332 | 1.5374 | 5.82E-01 | 7.53E-01 |
| ADA+ vs. ADA- | DQB1*06:02 | 1.2329 | 0.5612 | 2.7084 | 1.4941 | 6.02E-01 | 7.64E-01 |
| ADA+ vs. ADA- | C*01:02 | 1.1732 | 0.6233 | 2.2082 | 1.3808 | 6.21E-01 | 7.65E-01 |
| ADA+ vs. ADA- | DQB1*03:01 | 0.8966 | 0.5760 | 1.3956 | 1.2533 | 6.29E-01 | 7.65E-01 |
| ADA+ vs. ADA- | DRA*01:01 | 0.8694 | 0.4858 | 1.5558 | 1.3457 | 6.37E-01 | 7.65E-01 |
| ADA+ vs. ADA- | DQB1*02:02 | 1.1010 | 0.6702 | 1.8087 | 1.2882 | 7.04E-01 | 8.30E-01 |
| ADA+ vs. ADA- | DQB1*03:03 | 1.1520 | 0.5072 | 2.6168 | 1.5198 | 7.35E-01 | 8.39E-01 |
| ADA+ vs. ADA- | C*06:02 | 0.8924 | 0.4587 | 1.7363 | 1.4044 | 7.37E-01 | 8.39E-01 |
| ADA+ vs. ADA- | C*04:01 | 0.9225 | 0.5503 | 1.5465 | 1.3016 | 7.60E-01 | 8.41E-01 |
| ADA+ vs. ADA- | B*13:02 | 1.1097 | 0.5614 | 2.1935 | 1.4158 | 7.65E-01 | 8.41E-01 |
| ADA+ vs. ADA- | DQA1*03:01 | 1.1285 | 0.4636 | 2.7472 | 1.5745 | 7.90E-01 | 8.55E-01 |
| ADA+ vs. ADA- | B*27:05 | 1.0836 | 0.4804 | 2.4438 | 1.5143 | 8.47E-01 | 9.01E-01 |
| ADA+ vs. ADA- | B*07:02 | 1.0369 | 0.5772 | 1.8626 | 1.3483 | 9.04E-01 | 9.47E-01 |
| ADA+ vs. ADA- | DQB1*05:02 | 0.9386 | 0.2529 | 3.4830 | 1.9524 | 9.25E-01 | 9.53E-01 |
| ADA+ vs. ADA- | DQB1*03:02 | 1.0419 | 0.3434 | 3.1616 | 1.7618 | 9.42E-01 | 9.57E-01 |

| | | | | | | | |
|---|---|---|---|---|---|---|---|
| ADA+ vs. ADA- | C*07:02 | 0.9982 | 0.4280 | 2.3283 | 1.5405 | 9.97E-01 | 9.97E-01 |
| NAb+ vs. ADA- | DQB1*06:03 | 13.0938 | 6.4793 | 26.4607 | 1.4318 | 7.73E-13 | 4.87E-11 |
| NAb+ vs. ADA- | DRB1*13:01 | 10.4178 | 4.8170 | 22.5308 | 1.4823 | 2.61E-09 | 8.21E-08 |
| NAb+ vs. ADA- | DQA1*01:03 | 6.1363 | 3.2242 | 11.6787 | 1.3887 | 3.29E-08 | 6.90E-07 |
| NAb+ vs. ADA- | C*03:03 | 3.1964 | 1.5304 | 6.6762 | 1.4561 | 1.99E-03 | 3.13E-02 |
| NAb+ vs. ADA- | B*15:01 | 2.8787 | 1.4391 | 5.7584 | 1.4244 | 2.80E-03 | 3.53E-02 |
| NAb+ vs. ADA- | DRB1*04:01 | 4.6700 | 1.3819 | 15.7818 | 1.8613 | 1.31E-02 | 1.38E-01 |
| NAb+ vs. ADA- | DQA1*03:03 | 3.6939 | 1.2629 | 10.8045 | 1.7291 | 1.70E-02 | 1.53E-01 |
| NAb+ vs. ADA- | C*03:04 | 2.5873 | 1.1315 | 5.9159 | 1.5250 | 2.43E-02 | 1.91E-01 |
| NAb+ vs. ADA- | C*05:01 | 2.2890 | 1.0796 | 4.8532 | 1.4673 | 3.08E-02 | 2.16E-01 |
| NAb+ vs. ADA- | DQB1*05:01 | 0.4847 | 0.2403 | 0.9777 | 1.4305 | 4.31E-02 | 2.71E-01 |
| NAb+ vs. ADA- | DQA1*03:01 | 2.0287 | 0.9832 | 4.1860 | 1.4471 | 5.56E-02 | 3.19E-01 |
| NAb+ vs. ADA- | A*25:01 | 2.6637 | 0.9513 | 7.4589 | 1.6911 | 6.22E-02 | 3.27E-01 |
| NAb+ vs. ADA- | DQA1*01:01 | 0.5296 | 0.2616 | 1.0721 | 1.4330 | 7.73E-02 | 3.75E-01 |
| NAb+ vs. ADA- | B*14:02 | 0.2499 | 0.0508 | 1.2294 | 2.2546 | 8.80E-02 | 3.87E-01 |
| NAb+ vs. ADA- | B*44:02 | 1.8433 | 0.9045 | 3.7565 | 1.4380 | 9.22E-02 | 3.87E-01 |
| NAb+ vs. ADA- | C*08:02 | 0.2716 | 0.0569 | 1.2967 | 2.2203 | 1.02E-01 | 3.93E-01 |
| NAb+ vs. ADA- | DRB1*11:04 | 0.3671 | 0.1071 | 1.2582 | 1.8748 | 1.11E-01 | 3.93E-01 |
| NAb+ vs. ADA- | DQB1*02:01 | 0.5490 | 0.2577 | 1.1693 | 1.4708 | 1.20E-01 | 3.93E-01 |
| NAb+ vs. ADA- | B*08:01 | 0.5448 | 0.2510 | 1.1826 | 1.4850 | 1.25E-01 | 3.93E-01 |
| NAb+ vs. ADA- | DRB1*03:01 | 0.5561 | 0.2612 | 1.1842 | 1.4705 | 1.28E-01 | 3.93E-01 |
| NAb+ vs. ADA- | C*02:02 | 0.3175 | 0.0717 | 1.4064 | 2.1368 | 1.31E-01 | 3.93E-01 |
| NAb+ vs. ADA- | C*06:02 | 0.5279 | 0.2210 | 1.2609 | 1.5593 | 1.50E-01 | 4.31E-01 |
| NAb+ vs. ADA- | DRB1*11:01 | 0.5822 | 0.2602 | 1.3026 | 1.5082 | 1.88E-01 | 4.80E-01 |
| NAb+ vs. ADA- | DQA1*05:01 | 0.6117 | 0.2941 | 1.2722 | 1.4529 | 1.88E-01 | 4.80E-01 |
| NAb+ vs. ADA- | DQA1*05:05 | 0.5091 | 0.1854 | 1.3986 | 1.6746 | 1.90E-01 | 4.80E-01 |
| NAb+ vs. ADA- | DPA1*02:01 | 1.4886 | 0.7839 | 2.8268 | 1.3871 | 2.24E-01 | 5.27E-01 |
| NAb+ vs. ADA- | C*12:03 | 2.0621 | 0.6067 | 7.0088 | 1.8668 | 2.46E-01 | 5.27E-01 |
| NAb+ vs. ADA- | B*40:01 | 1.7940 | 0.6589 | 4.8844 | 1.6670 | 2.53E-01 | 5.27E-01 |
| NAb+ vs. ADA- | A*01:01 | 0.7138 | 0.3964 | 1.2853 | 1.3500 | 2.61E-01 | 5.27E-01 |
| NAb+ vs. ADA- | DQB1*03:02 | 1.7300 | 0.6637 | 4.5093 | 1.6303 | 2.62E-01 | 5.27E-01 |
| NAb+ vs. ADA- | DQA1*01:04 | 0.4285 | 0.0959 | 1.9144 | 2.1463 | 2.67E-01 | 5.27E-01 |
| NAb+ vs. ADA- | A*68:01 | 2.3850 | 0.5132 | 11.0839 | 2.1899 | 2.67E-01 | 5.27E-01 |
| NAb+ vs. ADA- | DQB1*05:03 | 0.4424 | 0.0979 | 1.9995 | 2.1589 | 2.89E-01 | 5.52E-01 |
| NAb+ vs. ADA- | DPA1*01:03 | 0.5378 | 0.1510 | 1.9156 | 1.9119 | 3.39E-01 | 6.27E-01 |

| | | | | | | | |
|---|---|---|---|---|---|---|---|
| NAb+ vs. ADA- | B*18:01 | 1.4394 | 0.6460 | 3.2071 | 1.5050 | 3.73E-01 | 6.42E-01 |
| NAb+ vs. ADA- | DRA*01:01 | 0.7321 | 0.3673 | 1.4590 | 1.4217 | 3.75E-01 | 6.42E-01 |
| NAb+ vs. ADA- | DRA*01:02 | 1.2696 | 0.7449 | 2.1640 | 1.3127 | 3.80E-01 | 6.42E-01 |
| NAb+ vs. ADA- | A*02:01 | 1.2557 | 0.7493 | 2.1041 | 1.3013 | 3.87E-01 | 6.42E-01 |
| NAb+ vs. ADA- | DRB1*14:54 | 0.4179 | 0.0529 | 3.3001 | 2.8701 | 4.08E-01 | 6.59E-01 |
| NAb+ vs. ADA- | DRB1*12:01 | 1.5260 | 0.4825 | 4.8261 | 1.7994 | 4.72E-01 | 7.43E-01 |
| NAb+ vs. ADA- | DRB1*01:01 | 0.7520 | 0.3187 | 1.7739 | 1.5495 | 5.15E-01 | 7.60E-01 |
| NAb+ vs. ADA- | B*51:01 | 1.3381 | 0.5479 | 3.2677 | 1.5770 | 5.23E-01 | 7.60E-01 |
| NAb+ vs. ADA- | A*03:01 | 1.3512 | 0.5332 | 3.4239 | 1.6070 | 5.26E-01 | 7.60E-01 |
| NAb+ vs. ADA- | DRB1*15:01 | 1.3103 | 0.5627 | 3.0510 | 1.5392 | 5.31E-01 | 7.60E-01 |
| NAb+ vs. ADA- | C*07:02 | 1.2963 | 0.5249 | 3.2014 | 1.5861 | 5.74E-01 | 7.64E-01 |
| NAb+ vs. ADA- | C*01:02 | 0.7722 | 0.3086 | 1.9323 | 1.5967 | 5.81E-01 | 7.64E-01 |
| NAb+ vs. ADA- | B*27:05 | 0.7622 | 0.2833 | 2.0508 | 1.6570 | 5.91E-01 | 7.64E-01 |
| NAb+ vs. ADA- | C*04:01 | 1.1724 | 0.6484 | 2.1196 | 1.3528 | 5.99E-01 | 7.64E-01 |
| NAb+ vs. ADA- | DQA1*02:01 | 0.8283 | 0.4047 | 1.6950 | 1.4410 | 6.06E-01 | 7.64E-01 |
| NAb+ vs. ADA- | DRB1*07:01 | 0.8283 | 0.4047 | 1.6950 | 1.4410 | 6.06E-01 | 7.64E-01 |
| NAb+ vs. ADA- | DQB1*02:02 | 0.8271 | 0.3825 | 1.7885 | 1.4821 | 6.30E-01 | 7.78E-01 |
| NAb+ vs. ADA- | B*07:02 | 1.1446 | 0.5555 | 2.3586 | 1.4461 | 7.14E-01 | 8.65E-01 |
| NAb+ vs. ADA- | DQB1*03:01 | 0.8769 | 0.3903 | 1.9700 | 1.5113 | 7.50E-01 | 8.92E-01 |
| NAb+ vs. ADA- | C*07:01 | 0.9247 | 0.5276 | 1.6207 | 1.3315 | 7.85E-01 | 9.05E-01 |
| NAb+ vs. ADA- | DRB1*13:02 | 1.1539 | 0.3749 | 3.5513 | 1.7746 | 8.03E-01 | 9.05E-01 |
| NAb+ vs. ADA- | B*13:02 | 0.8674 | 0.2755 | 2.7307 | 1.7952 | 8.08E-01 | 9.05E-01 |
| NAb+ vs. ADA- | DQB1*06:02 | 1.1203 | 0.4234 | 2.9638 | 1.6428 | 8.19E-01 | 9.05E-01 |
| NAb+ vs. ADA- | B*35:03 | 1.1092 | 0.3892 | 3.1605 | 1.7062 | 8.46E-01 | 9.19E-01 |
| NAb+ vs. ADA- | B*52:01 | 1.1132 | 0.3060 | 4.0499 | 1.9327 | 8.71E-01 | 9.20E-01 |
| NAb+ vs. ADA- | A*24:02 | 0.9411 | 0.4380 | 2.0221 | 1.4773 | 8.76E-01 | 9.20E-01 |
| NAb+ vs. ADA- | A*32:01 | 0.9450 | 0.2652 | 3.3675 | 1.9124 | 9.30E-01 | 9.52E-01 |
| NAb+ vs. ADA- | DQA1*01:02 | 0.9763 | 0.5105 | 1.8672 | 1.3921 | 9.42E-01 | 9.52E-01 |
| NAb+ vs. ADA- | B*35:01 | 1.0321 | 0.3718 | 2.8651 | 1.6836 | 9.52E-01 | 9.52E-01 |

***Supplemental Table 2: Conditional meta-analysis results (conditioned on DQB1*06:03)***

| Analysis | allele | OR | conf.lower | conf.upper | seTE | pval | pval.bh |
|---|---|---|---|---|---|---|---|
| ADA+ vs. ADA- | DQA1*03:03 | 2.8294 | 1.5203 | 5.2656 | 1.3729 | 1.03E-03 | 3.75E-02 |
| ADA+ vs. ADA- | DQA1*05:01 | 0.3671 | 0.1977 | 0.6815 | 1.3712 | 1.50E-03 | 3.75E-02 |
| ADA+ vs. ADA- | DRB1*03:01 | 0.3622 | 0.1904 | 0.6889 | 1.3882 | 1.96E-03 | 3.75E-02 |
| ADA+ vs. ADA- | DQB1*02:01 | 0.3635 | 0.1896 | 0.6969 | 1.3938 | 2.31E-03 | 3.75E-02 |
| ADA+ vs. ADA- | DRB1*04:01 | 3.3228 | 1.4439 | 7.6466 | 1.5299 | 4.74E-03 | 5.50E-02 |
| ADA+ vs. ADA- | A*01:01 | 0.5115 | 0.3200 | 0.8175 | 1.2703 | 5.08E-03 | 5.50E-02 |
| ADA+ vs. ADA- | C*03:04 | 2.4346 | 1.2746 | 4.6503 | 1.3912 | 7.04E-03 | 6.54E-02 |
| ADA+ vs. ADA- | C*07:01 | 0.5814 | 0.3699 | 0.9136 | 1.2594 | 1.87E-02 | 1.52E-01 |
| ADA+ vs. ADA- | A*68:01 | 2.4088 | 1.1328 | 5.1219 | 1.4695 | 2.24E-02 | 1.62E-01 |
| ADA+ vs. ADA- | A*26:01 | 2.2459 | 1.0829 | 4.6579 | 1.4509 | 2.97E-02 | 1.93E-01 |
| ADA+ vs. ADA- | B*15:01 | 1.8433 | 1.0294 | 3.3006 | 1.3461 | 3.96E-02 | 2.21E-01 |
| ADA+ vs. ADA- | B*08:01 | 0.4175 | 0.1807 | 0.9644 | 1.5330 | 4.09E-02 | 2.21E-01 |
| ADA+ vs. ADA- | DQA1*01:02 | 1.5198 | 0.9395 | 2.4586 | 1.2782 | 8.81E-02 | 4.41E-01 |
| ADA+ vs. ADA- | DRB1*15:01 | 1.6542 | 0.9028 | 3.0308 | 1.3620 | 1.03E-01 | 4.52E-01 |
| ADA+ vs. ADA- | DPA1*02:01 | 1.4203 | 0.9236 | 2.1841 | 1.2455 | 1.10E-01 | 4.52E-01 |
| ADA+ vs. ADA- | B*40:01 | 1.7516 | 0.8786 | 3.4919 | 1.4219 | 1.11E-01 | 4.52E-01 |
| ADA+ vs. ADA- | DQA1*02:01 | 1.4464 | 0.8950 | 2.3374 | 1.2775 | 1.32E-01 | 4.76E-01 |
| ADA+ vs. ADA- | DRB1*07:01 | 1.4464 | 0.8950 | 2.3374 | 1.2775 | 1.32E-01 | 4.76E-01 |
| ADA+ vs. ADA- | DQA1*05:05 | 0.6686 | 0.3691 | 1.2109 | 1.3540 | 1.84E-01 | 6.30E-01 |
| ADA+ vs. ADA- | A*02:01 | 1.2741 | 0.8658 | 1.8748 | 1.2179 | 2.19E-01 | 6.40E-01 |
| ADA+ vs. ADA- | DQA1*01:01 | 0.7455 | 0.4608 | 1.2062 | 1.2782 | 2.32E-01 | 6.40E-01 |
| ADA+ vs. ADA- | DQB1*05:01 | 0.7516 | 0.4702 | 1.2014 | 1.2703 | 2.33E-01 | 6.40E-01 |
| ADA+ vs. ADA- | DQB1*06:02 | 1.5759 | 0.7424 | 3.3456 | 1.4683 | 2.36E-01 | 6.40E-01 |
| ADA+ vs. ADA- | DQA1*01:03 | 0.5537 | 0.2063 | 1.4866 | 1.6551 | 2.41E-01 | 6.40E-01 |
| ADA+ vs. ADA- | DQB1*02:02 | 1.3469 | 0.7997 | 2.2686 | 1.3047 | 2.63E-01 | 6.40E-01 |
| ADA+ vs. ADA- | C*12:03 | 1.7109 | 0.6647 | 4.4039 | 1.6199 | 2.66E-01 | 6.40E-01 |
| ADA+ vs. ADA- | DRB1*13:02 | 1.5698 | 0.6951 | 3.5449 | 1.5153 | 2.78E-01 | 6.40E-01 |
| ADA+ vs. ADA- | DRB1*11:04 | 0.6863 | 0.3468 | 1.3581 | 1.4166 | 2.80E-01 | 6.40E-01 |
| ADA+ vs. ADA- | DRB1*11:01 | 0.7278 | 0.4062 | 1.3038 | 1.3465 | 2.85E-01 | 6.40E-01 |
| ADA+ vs. ADA- | B*51:01 | 1.3761 | 0.7526 | 2.5162 | 1.3606 | 3.00E-01 | 6.50E-01 |
| ADA+ vs. ADA- | DQA1*03:01 | 1.5351 | 0.6579 | 3.5823 | 1.5409 | 3.21E-01 | 6.70E-01 |
| ADA+ vs. ADA- | C*05:01 | 1.3321 | 0.7481 | 2.3722 | 1.3423 | 3.30E-01 | 6.70E-01 |
| ADA+ vs. ADA- | A*25:01 | 1.4643 | 0.6510 | 3.2936 | 1.5122 | 3.56E-01 | 7.02E-01 |

| | | | | | | | |
|---|---|---|---|---|---|---|---|
| ADA+ vs. ADA- | DPA1*01:03 | 0.6475 | 0.2358 | 1.7779 | 1.6743 | 3.99E-01 | 7.48E-01 |
| ADA+ vs. ADA- | B*52:01 | 0.6610 | 0.2438 | 1.7922 | 1.6635 | 4.16E-01 | 7.48E-01 |
| ADA+ vs. ADA- | DQB1*03:03 | 1.4117 | 0.6078 | 3.2788 | 1.5372 | 4.23E-01 | 7.48E-01 |
| ADA+ vs. ADA- | B*27:05 | 1.3194 | 0.6629 | 2.6259 | 1.4207 | 4.30E-01 | 7.48E-01 |
| ADA+ vs. ADA- | DRA*01:02 | 1.1721 | 0.7847 | 1.7507 | 1.2272 | 4.38E-01 | 7.48E-01 |
| ADA+ vs. ADA- | A*24:02 | 0.8079 | 0.4653 | 1.4027 | 1.3251 | 4.49E-01 | 7.48E-01 |
| ADA+ vs. ADA- | C*02:02 | 0.7072 | 0.2739 | 1.8264 | 1.6226 | 4.74E-01 | 7.49E-01 |
| ADA+ vs. ADA- | B*44:02 | 1.2328 | 0.6898 | 2.2032 | 1.3448 | 4.80E-01 | 7.49E-01 |
| ADA+ vs. ADA- | A*32:01 | 0.7770 | 0.3760 | 1.6054 | 1.4481 | 4.96E-01 | 7.49E-01 |
| ADA+ vs. ADA- | B*14:02 | 0.7268 | 0.2805 | 1.8830 | 1.6253 | 5.11E-01 | 7.49E-01 |
| ADA+ vs. ADA- | A*03:01 | 1.1949 | 0.6979 | 2.0459 | 1.3157 | 5.16E-01 | 7.49E-01 |
| ADA+ vs. ADA- | DQB1*03:02 | 1.4052 | 0.5003 | 3.9468 | 1.6937 | 5.19E-01 | 7.49E-01 |
| ADA+ vs. ADA- | C*08:02 | 0.7636 | 0.3147 | 1.8530 | 1.5719 | 5.51E-01 | 7.79E-01 |
| ADA+ vs. ADA- | DQB1*05:03 | 0.8265 | 0.3860 | 1.7696 | 1.4747 | 6.24E-01 | 8.36E-01 |
| ADA+ vs. ADA- | DQA1*01:04 | 0.8395 | 0.3935 | 1.7910 | 1.4719 | 6.51E-01 | 8.36E-01 |
| ADA+ vs. ADA- | B*35:03 | 0.8164 | 0.3284 | 2.0300 | 1.5916 | 6.63E-01 | 8.36E-01 |
| ADA+ vs. ADA- | DRB1*13:01 | 0.7391 | 0.1860 | 2.9360 | 2.0214 | 6.67E-01 | 8.36E-01 |
| ADA+ vs. ADA- | B*13:02 | 1.1977 | 0.5188 | 2.7653 | 1.5325 | 6.73E-01 | 8.36E-01 |
| ADA+ vs. ADA- | B*07:02 | 1.1108 | 0.6752 | 1.8274 | 1.2892 | 6.79E-01 | 8.36E-01 |
| ADA+ vs. ADA- | DQB1*03:01 | 1.0847 | 0.7356 | 1.5995 | 1.2192 | 6.82E-01 | 8.36E-01 |
| ADA+ vs. ADA- | B*38:01 | 1.3987 | 0.2483 | 7.8795 | 2.4158 | 7.04E-01 | 8.47E-01 |
| ADA+ vs. ADA- | B*18:01 | 1.1055 | 0.5711 | 2.1400 | 1.4007 | 7.66E-01 | 8.96E-01 |
| ADA+ vs. ADA- | B*35:01 | 0.8546 | 0.2916 | 2.5046 | 1.7308 | 7.75E-01 | 8.96E-01 |
| ADA+ vs. ADA- | C*03:03 | 1.0992 | 0.5564 | 2.1716 | 1.4153 | 7.85E-01 | 8.96E-01 |
| ADA+ vs. ADA- | C*01:02 | 1.0854 | 0.5624 | 2.0951 | 1.3987 | 8.07E-01 | 8.97E-01 |
| ADA+ vs. ADA- | DRB1*14:54 | 0.9079 | 0.4051 | 2.0349 | 1.5095 | 8.14E-01 | 8.97E-01 |
| ADA+ vs. ADA- | DRB1*01:01 | 0.9416 | 0.5408 | 1.6392 | 1.3269 | 8.31E-01 | 9.01E-01 |
| ADA+ vs. ADA- | DRA*01:01 | 1.0426 | 0.6335 | 1.7159 | 1.2894 | 8.70E-01 | 9.19E-01 |
| ADA+ vs. ADA- | C*07:02 | 1.0610 | 0.5018 | 2.2431 | 1.4652 | 8.77E-01 | 9.19E-01 |
| ADA+ vs. ADA- | C*06:02 | 0.9847 | 0.5393 | 1.7982 | 1.3596 | 9.60E-01 | 9.81E-01 |
| ADA+ vs. ADA- | C*04:01 | 0.9859 | 0.5117 | 1.8998 | 1.3975 | 9.66E-01 | 9.81E-01 |
| ADA+ vs. ADA- | DQB1*05:02 | 0.9940 | 0.2189 | 4.5136 | 2.1641 | 9.94E-01 | 9.94E-01 |
| NAb+ vs. ADA- | DQA1*03:03 | 4.9795 | 1.8664 | 13.2852 | 1.6498 | 1.34E-03 | 4.41E-02 |
| NAb+ vs. ADA- | DQA1*03:01 | 3.5068 | 1.6223 | 7.5805 | 1.4819 | 1.42E-03 | 4.41E-02 |
| NAb+ vs. ADA- | DRB1*04:01 | 6.7646 | 1.9617 | 23.3263 | 1.8806 | 2.47E-03 | 4.45E-02 |

| | | | | | | | |
|---|---|---|---|---|---|---|---|
| NAb+ vs. ADA- | C*03:04 | 4.0183 | 1.6105 | 10.0260 | 1.5944 | 2.87E-03 | 4.45E-02 |
| NAb+ vs. ADA- | B*15:01 | 2.3350 | 1.0759 | 5.0676 | 1.4849 | 3.20E-02 | 3.96E-01 |
| NAb+ vs. ADA- | DQB1*03:02 | 2.6762 | 1.0106 | 7.0870 | 1.6436 | 4.76E-02 | 4.92E-01 |
| NAb+ vs. ADA- | B*44:02 | 2.1263 | 0.9729 | 4.6470 | 1.4902 | 5.86E-02 | 5.19E-01 |
| NAb+ vs. ADA- | A*25:01 | 2.7374 | 0.9267 | 8.0855 | 1.7378 | 6.84E-02 | 5.30E-01 |
| NAb+ vs. ADA- | DQB1*02:01 | 0.4757 | 0.2044 | 1.1071 | 1.5388 | 8.47E-02 | 5.38E-01 |
| NAb+ vs. ADA- | C*05:01 | 2.3043 | 0.8444 | 6.2880 | 1.6689 | 1.03E-01 | 5.38E-01 |
| NAb+ vs. ADA- | B*08:01 | 0.4982 | 0.2133 | 1.1634 | 1.5415 | 1.07E-01 | 5.38E-01 |
| NAb+ vs. ADA- | C*03:03 | 1.9733 | 0.8375 | 4.6499 | 1.5485 | 1.20E-01 | 5.38E-01 |
| NAb+ vs. ADA- | DRB1*03:01 | 0.5161 | 0.2233 | 1.1929 | 1.5334 | 1.22E-01 | 5.38E-01 |
| NAb+ vs. ADA- | DQA1*05:01 | 0.5223 | 0.2263 | 1.2056 | 1.5324 | 1.28E-01 | 5.38E-01 |
| NAb+ vs. ADA- | A*01:01 | 0.6083 | 0.3151 | 1.1745 | 1.3989 | 1.39E-01 | 5.38E-01 |
| NAb+ vs. ADA- | DRB1*11:04 | 0.3834 | 0.1053 | 1.3958 | 1.9332 | 1.46E-01 | 5.38E-01 |
| NAb+ vs. ADA- | B*40:01 | 2.2142 | 0.7404 | 6.6216 | 1.7488 | 1.55E-01 | 5.38E-01 |
| NAb+ vs. ADA- | C*06:02 | 0.4960 | 0.1881 | 1.3075 | 1.6398 | 1.56E-01 | 5.38E-01 |
| NAb+ vs. ADA- | C*02:02 | 0.3409 | 0.0716 | 1.6242 | 2.2177 | 1.77E-01 | 5.77E-01 |
| NAb+ vs. ADA- | A*02:01 | 1.3974 | 0.7894 | 2.4737 | 1.3383 | 2.51E-01 | 7.42E-01 |
| NAb+ vs. ADA- | DQA1*05:05 | 0.5718 | 0.2151 | 1.5198 | 1.6467 | 2.62E-01 | 7.42E-01 |
| NAb+ vs. ADA- | B*14:02 | 0.3891 | 0.0744 | 2.0338 | 2.3251 | 2.63E-01 | 7.42E-01 |
| NAb+ vs. ADA- | C*08:02 | 0.4102 | 0.0803 | 2.0953 | 2.2981 | 2.84E-01 | 7.66E-01 |
| NAb+ vs. ADA- | DRB1*13:02 | 1.7933 | 0.5606 | 5.7370 | 1.8099 | 3.25E-01 | 7.80E-01 |
| NAb+ vs. ADA- | A*68:01 | 2.1648 | 0.4284 | 10.9388 | 2.2854 | 3.50E-01 | 7.80E-01 |
| NAb+ vs. ADA- | DPA1*02:01 | 1.3990 | 0.6806 | 2.8756 | 1.4443 | 3.61E-01 | 7.80E-01 |
| NAb+ vs. ADA- | DQB1*06:02 | 1.4322 | 0.6553 | 3.1302 | 1.4902 | 3.68E-01 | 7.80E-01 |
| NAb+ vs. ADA- | DRB1*12:01 | 1.7957 | 0.5008 | 6.4389 | 1.9184 | 3.69E-01 | 7.80E-01 |
| NAb+ vs. ADA- | DRB1*15:01 | 1.5350 | 0.5937 | 3.9690 | 1.6236 | 3.77E-01 | 7.80E-01 |
| NAb+ vs. ADA- | DRB1*14:54 | 0.3538 | 0.0344 | 3.6363 | 3.2829 | 3.82E-01 | 7.80E-01 |
| NAb+ vs. ADA- | DRB1*11:01 | 0.6873 | 0.2882 | 1.6391 | 1.5580 | 3.98E-01 | 7.80E-01 |
| NAb+ vs. ADA- | DQB1*05:01 | 0.7404 | 0.3560 | 1.5400 | 1.4530 | 4.21E-01 | 7.80E-01 |
| NAb+ vs. ADA- | C*01:02 | 0.6646 | 0.2411 | 1.8321 | 1.6776 | 4.30E-01 | 7.80E-01 |
| NAb+ vs. ADA- | C*04:01 | 1.3035 | 0.6745 | 2.5191 | 1.3995 | 4.30E-01 | 7.80E-01 |
| NAb+ vs. ADA- | DQA1*01:04 | 0.5396 | 0.1074 | 2.7118 | 2.2791 | 4.54E-01 | 7.80E-01 |
| NAb+ vs. ADA- | C*07:01 | 0.7909 | 0.4249 | 1.4720 | 1.3729 | 4.59E-01 | 7.80E-01 |
| NAb+ vs. ADA- | C*07:02 | 1.2602 | 0.6638 | 2.3926 | 1.3870 | 4.80E-01 | 7.80E-01 |
| NAb+ vs. ADA- | DQB1*05:03 | 0.5632 | 0.1107 | 2.8668 | 2.2939 | 4.89E-01 | 7.80E-01 |

| | | | | | | | |
|---|---|---|---|---|---|---|---|
| NAb+ vs. ADA- | B*52:01 | 1.6014 | 0.4068 | 6.3040 | 2.0120 | 5.01E-01 | 7.80E-01 |
| NAb+ vs. ADA- | B*18:01 | 1.3510 | 0.5601 | 3.2589 | 1.5671 | 5.03E-01 | 7.80E-01 |
| NAb+ vs. ADA- | B*35:01 | 1.4332 | 0.4812 | 4.2685 | 1.7451 | 5.18E-01 | 7.83E-01 |
| NAb+ vs. ADA- | B*07:02 | 1.2088 | 0.6256 | 2.3355 | 1.3994 | 5.73E-01 | 8.30E-01 |
| NAb+ vs. ADA- | DQA1*01:01 | 0.8105 | 0.3885 | 1.6910 | 1.4553 | 5.75E-01 | 8.30E-01 |
| NAb+ vs. ADA- | DRB1*13:01 | 0.6897 | 0.1370 | 3.4731 | 2.2813 | 6.52E-01 | 9.07E-01 |
| NAb+ vs. ADA- | A*03:01 | 1.2424 | 0.4750 | 3.2495 | 1.6332 | 6.58E-01 | 9.07E-01 |
| NAb+ vs. ADA- | DPA1*01:03 | 0.7732 | 0.1831 | 3.2651 | 2.0854 | 7.26E-01 | 9.39E-01 |
| NAb+ vs. ADA- | DQA1*01:02 | 1.1805 | 0.4331 | 3.2180 | 1.6681 | 7.46E-01 | 9.39E-01 |
| NAb+ vs. ADA- | A*24:02 | 0.8686 | 0.3613 | 2.0882 | 1.5644 | 7.53E-01 | 9.39E-01 |
| NAb+ vs. ADA- | B*51:01 | 1.1626 | 0.4449 | 3.0383 | 1.6325 | 7.59E-01 | 9.39E-01 |
| NAb+ vs. ADA- | DRA*01:02 | 1.0950 | 0.6076 | 1.9732 | 1.3505 | 7.63E-01 | 9.39E-01 |
| NAb+ vs. ADA- | C*12:03 | 1.1754 | 0.3802 | 3.6343 | 1.7788 | 7.79E-01 | 9.39E-01 |
| NAb+ vs. ADA- | B*13:02 | 0.8364 | 0.2277 | 3.0726 | 1.9423 | 7.88E-01 | 9.39E-01 |
| NAb+ vs. ADA- | DQA1*01:03 | 0.8528 | 0.2069 | 3.5153 | 2.0598 | 8.26E-01 | 9.48E-01 |
| NAb+ vs. ADA- | DRB1*01:01 | 1.0999 | 0.4710 | 2.5685 | 1.5414 | 8.26E-01 | 9.48E-01 |
| NAb+ vs. ADA- | DQB1*03:01 | 1.0797 | 0.4201 | 2.7750 | 1.6187 | 8.73E-01 | 9.49E-01 |
| NAb+ vs. ADA- | DQA1*02:01 | 1.0586 | 0.4901 | 2.2865 | 1.4813 | 8.85E-01 | 9.49E-01 |
| NAb+ vs. ADA- | DRB1*07:01 | 1.0586 | 0.4901 | 2.2865 | 1.4813 | 8.85E-01 | 9.49E-01 |
| NAb+ vs. ADA- | DRA*01:01 | 1.0532 | 0.5113 | 2.1694 | 1.4458 | 8.88E-01 | 9.49E-01 |
| NAb+ vs. ADA- | DQB1*02:02 | 1.0340 | 0.4538 | 2.3559 | 1.5222 | 9.37E-01 | 9.79E-01 |
| NAb+ vs. ADA- | B*35:03 | 1.0371 | 0.3091 | 3.4791 | 1.8544 | 9.53E-01 | 9.79E-01 |
| NAb+ vs. ADA- | A*32:01 | 1.0318 | 0.2715 | 3.9214 | 1.9762 | 9.63E-01 | 9.79E-01 |
| NAb+ vs. ADA- | B*27:05 | 0.9918 | 0.3444 | 2.8557 | 1.7153 | 9.88E-01 | 9.88E-01 |

***Supplemental Table 3: List of independent ethics committees / institutional review boards (Hickory)***

| Country | Authority/Committee Name | Authority/Committee Address |
|---|---|---|
| Argentina | Comite de Etica del Hospital Provincial del Centenario | Urquiza 3101 Rosario Santa Fe S2002KDS |
| Argentina | Comite de Etica Independiente Consultorios Integrados | Italia 428 Rosario Santa Fe 2000 |
| Argentina | Comité Institucional de Revisión de Estudios de Investigación (CIREI) | Córdoba 4545 Mar del Plata Buenos Aires B7602CBM |
| Argentina | Comité Médico de Docencia e Investigación de la Fundación Estudios Clínicos | Italia 428 Rosario Santa Fe 2000 |
| Australia | Cabrini Hospital Malvern | 181-183 Wattletree Rd Malvern<br>Melbourne<br>Victoria<br>3144 |
| Australia | Cabrini Human Research Ethics Committee | 154 Wattletree Road Level 1<br>Malvern<br>Victoria<br>3144 |
| Australia | Mater Health Services Human Research Ethics Committee | Annerley Road Level 3, Room 55-57<br>Quarters Building Woolloongabba<br>Queensland<br>4101 |
| Australia | Melbourne Health Human Research Ethics Committee (RGO) | Melbourne Health<br>Office for Research, Level 2 South West<br>Royal Melbourne Hospital, 300- 336<br>Grattan Street<br>Parkville<br>Victoria<br>3050 |
| Australia | Metro South Health Service District Human Research Ethics Committee (RGO) | Centres for Health Research Level 2,<br>Building 35<br>Princess Alexandra Hospital, Ispwich<br>Road Woolloongabba Queensland<br>4102 |
| Australia | Royal Brisbane & Women's Hospital RGO | Butterfield St Herston Queensland 4029 |

| | | |
|---|---|---|
| Australia | South Metropolitan Health Service Human Research Ethics Committee (RGO) | 189 Royal Street East Perth Western Australia 6004 |
| Australia | South Western Sydney Local Health District (RGO) | Locked Bag 7017 New South Wales New South Wales 1871 |
| Australia | Southern Adelaide Clinical Human Research Ethics Committee-RGO | Human Research Ethics Room 2A 221 Flinders Medical Centre Level 2 BEDFORD PARK<br>South Australia 5042 |
| Australia | St Vincent's Hospital (Melbourne) HREC D (RGO) | 27 Victoria Parade<br>level 5, Mary Aikenhead Building<br>Research Governance Unit Fitzroy Victoria<br>3065 |
| Australia | Sydney Local Health Network Human Research Ethics Committee - Concord (RGO) | Concord Repatriation General Hospital<br>Hospital Road<br>Concord<br>New South Wales 2139 |
| Australia | Tasmania Health & Medical Human Research Ethics Committee | 301 Sandy Bay Road<br>Sandy Bay Campus, Office of Research Services<br>University of Tasmania, Private Bag 01<br>Hobart<br>Tasmania<br>7001 |
| Belgium | AZ Delta | Westlaan 123 Roeselare 8800 |
| Belgium | CHU de Liège | Domaine Universitaire du Sart Tilman<br>Batiment B 35<br>Liege<br>B-4000 |
| Belgium | Comité d’éthique hospitalo- Facultaire Cliniques Universitaires Saint Luc | Promenade de l’Alma 51 bte B1.43.03<br>Bruxelles<br>1200 |
| Belgium | Comité Local d'Ethique Hospitalier - C.H.U. St.-Pierre | Bâtiment Direction Rue Haute 322<br>Bruxelles<br>1000 |
| Belgium | Ethisch Comité Universitair Ziekenhuis Gent | De Pintelaan 185<br>UZ Gent - Ethisch comite 1K4 Ghent<br>B-9000 |

| | | |
|---|---|---|
| Belgium | Ethische Commissie - Algemeen Ziekenhuis Sint- Elisabeth Herentals | Nederrij 133 Herentals 2200 |
| Belgium | GZA Ziekenhuizen | Oosterveldlaan 24 Wilrijk<br>2610 |
| Belgium | Imeldaziekenhuis | Imeldalaan 9 Bonheiden 2820 |
| Belgium | Universitair Ziekenhuis Brussel Commissie Medische Ethiek | Laarbeeklaan 101 Brussels<br>1090 |
| Belgium | UZ Leuven | Campus Gasthuisberg Herestraat 49<br>Leuven<br>3000 |
| Brazil | *Hospital*das* | Rua Ramiro Barcelos, 2350 Porto Alegre<br>Rio Grande do Sul 90035-903 |
| Brazil | CEP Associação dos Funcionários Públicos do Rio Grande do Sul - Hospital Ernesto Dornelles | Avenida Ipiranga, 1801 11o andar - Sala 03 Porto Alegre<br>Rio Grande do Sul 90160-092 |
| Brazil | CEP da Faculdade de Jaguariúna (CEP-FAJ) | Rua Amazonas, 504 Jaguariúna<br>Sao Paulo 13820-000 |
| Brazil | CEP da Faculdade de Medicina de Botucatu - UNESP/SP | Distrito de Rubião Junior Botucatu<br>Sao Paulo<br>18618-970 |
| Brazil | CEP da Faculdade de Medicina do ABC/SP | Avenida Lauro Gomes, 2000 Vila Sacadura Cabral<br>Santo André Sao Paulo 09060-870 |
| Brazil | CEP da Faculdade Una de Uberlandia | Alameda Paulina Malgonari, 59<br>Uberlandia<br>Minas Gerais<br>38411-206 |
| Brazil | CEP da Universidade da Região de Joinville - UNIVILLE | Rua Paulo Malschitzki, 10 Joinville<br>Santa Catarina 89219-710 |
| Brazil | CEP da Universidade Federal de São Paulo / UNIFESP / EPM | Rua Botucatu, 572 - Conjunto 14 - 1o andar<br>Vila Clementino<br>São Paulo<br>Sao Paulo 04023-061 |
| Brazil | CEP do Hospital Allberto Rassi - HGG | Rua 246, 25 Coimbra Goiânia Goiás<br>74535-170 |

| | | |
|---|---|---|
| Brazil | CEP do Hospital de Clinicas da Universidade Federal do Parana - HCUFPR / PR | Rua General Carneiro, 181 Alto da Glória<br>Curitiba<br>Paraná<br>80060-900 |
| Brazil | CEP do Hospital Moinhos de Vento/ RS | Rua Ramiro Barcelos, 910<br>Sala 902 - Centro Clínico Ramiro - Moinhos de Vento<br>Porto Alegre<br>Rio Grande do Sul<br>90035-001 |
| Brazil | CEP do Hospital Pró- Cardíaco/RJ | Rua Voluntários da Pátria, 435 - 8o andar<br>Botafogo<br>Rio de Janeiro<br>Rio do Janeiro 22270-005 |
| Brazil | CEP do Hospital Universitário Clementino Fraga Filho - UFRJ | Rua Rodolpho Paulo Rocco, 255 - 1o andar - Sala 1D46<br>Cidade Universitária - Ilha do Fundão<br>Rio do Janeiro 21941-913 |
| Brazil | CEP do Instituto de Cardiologia do Distrito Federal | Estrada Parque Contorno do Bosque S/N – Cruzeiro Novo Brasília<br>Distrito Federal<br>70658-700 |
| Canada | Capital Health Authority Research Ethics Boards | 5790 University Avenue Room 240<br>Halifax<br>Nova Scotia<br>B3H 1V7 |
| Canada | Comité d'éthique de la recherche | 143 Wolfe Street Levis<br>Quebec<br>G6V 3Z1 |
| Canada | Conjoint Health Research Ethics Board | Room 9, Heritage Medical Research Buildig<br>Faculty of Medicine University of Calgary<br>Calgary<br>Alberta T2N 4N1 |
| Canada | Jewish General Hospital Research Ethics Office | 3755 cote ste-catherine Montreal<br>Quebec<br>H3T 1E2 |

| | | |
|---|---|---|
| Canada | Maisonneuve-Rosemont hospital REB | 5415 l'Assomption Blvd Montreal<br>Quebec<br>H1T 2M4 |
| Canada | Mount Sinai Hospital | 600 University Ave Infectious Control<br>Research, Room 171<br>Toronto<br>ON<br>M5G 1X5 |
| Canada | Mount Sinai Hospital Research Ethics Board | 700 University Avenue Suite 8-600<br>Toronto<br>Ontario<br>M5G 1Z5 |
| Canada | University of Alberta Health Research Ethics Board | 308 Campus Tower 8625 – 112 St.<br>Edmonton<br>Alberta<br>T6G 1K8 |
| Canada | University of Manitoba Health Services | 770 Bannatyne Avenue P126 Pathology<br>Building Winnipeg<br>Manitoba<br>R3E 0W3 |
| Canada | University of Saskatchewan Biomedical | Box 5000 RPO University Saskatoon<br>Saskatchewan<br>S7N 4J8 |
| Canada | University of Western Ontario | The University of Western Ontario<br>Office of Research<br>1151 Richmond Street London<br>Ontario N6A 3K7 |
| Canada | WIRB | 1019 39th Avenue SE Suite 120<br>Puyallup Washington 98374 |
| Czech Republic | Eticka komise - Krajska zdravotni a.s. | Socialni pece 3316/12A Usti nad Labem<br>40113 |
| Czech Republic | Eticka komise Fakultni nemocnice Hradec Kralove | Sokolska 581 Hradec Kralove 50005 |
| Czech Republic | Eticka komise Fakultni nemocnice u sv. Anny v Brne | Pekarska 53 Brno<br>656 91 |
| Czech Republic | Eticka komise FN a LF UP Olomouc | I.P.Pavlova 6 Olomouc 775 20 |

| | | |
|---|---|---|
| Czech Republic | Eticka komise IKEM a FTNsP | Videnska 800 Praha 4 - Krc 140 59 |
| Czech Republic | Eticka komise ISCARE I.V.F a.s. | Jankovcova 1569/2c Praha 7<br>17004 |
| Czech Republic | Eticka komise Nemocnice Na Bulovce | Budinova 67/2<br>Nemocnice Na Bulovce<br>Gynekologicko-porodnicka klinika<br>Praha 8-Liben<br>180 81 |
| Czech Republic | Eticka komise Pardubicke krajske nemocnice | Kyjevska 44 Pardubice 53301 |
| Greece | Scientific Committee of Anticancer Hospital of Thessaliniki " Theagenio" | 2 Alex. Symeonidi Str. Thessaloniki<br>54007 |
| Greece | Scientific Committee of General Hopsital of Athens "Laiko" | Mikras Asias Street Athens<br>11527 |
| Greece | Scientific Committee of General Hospital of Athens "Evangelismos" | 45-47 Ipsilandou Str Athens<br>10676 |
| Greece | Scientific Committee of General Hospital of Nea Ionia "Kostantopoulio" | 3-5 Agias Olgas Nea Ionia Athens<br>14233 |
| Greece | Scientific Committee of Interbalkan Hospital of Thessaloniki | 10, Asklipios Str. Pylaia Thessaloniki<br>57001 |
| Greece | Scientific Committee of University General Hospital of Alexandroupolis | Dragana Alexandroupolis 68100 |
| Greece | Scientific Committee of University General Hospital of Heraklion | Voutes P.O BOX 1352 Crete<br>71110 |
| Greece | Scientific Committee of University General Hospital of Ioannina | Stavros Niarchos Ave. Ioannina<br>45500 |
| Hungary | Egeszsegugyi Tudomanyos Tanacs Klinikai Farmakologiai Etikai Bizottsaga | Alkotmany u.25 Budapest<br>1054 |
| Hungary | Ethik-Kommission der Medizinischen Fakultaet der Ernst-Moritz-Arndt-Universitaet Greifswald | Felix-Hausdorff-Str. 3 Greifswald<br>Mecklenburg Vorpommern 17475 |
| Israel | Assaf Harofeh MC Ethics Committee | Zerifin<br>Assaf Harofeh Center Rishon Lezion<br>75141 |
| Israel | Chaim Sheba MC Ethics Committee | Tel Hashomer Ramat Gan 5265601 |

| | | |
|---|---|---|
| Israel | Hadassah MC Ethics Committee | Kiryat Hadassah POB 12000 Jerusalem<br>91120 |
| Israel | Kaplan MC Ethics Committee | Pasternak way St. POB 1<br>Rehovot<br>76100 |
| Israel | Rabin MC Ethics Committee | 39 Jabotinsky St Ground floor Beilinson Campus Raphael Recanati Genetic Inst. Blg Petach Tikva<br>4941492 |
| Israel | Rambam Health Care Campus Ethics Committee | 8 Haaliyya St., Bat Galim POB 9602<br>Haifa<br>3109601 |
| Israel | Shaare Zedek MC Ethics Committee | 12 Hans Bayt Bait Vagan Jerusalem<br>9103102 |
| Italy | Azienda Ospedaliera Universitaria Policlinico Tor Vergata | Viale Oxford, 81<br>Azienda Ospedaliera Univ Pol Tor Vergata - Dermatologia Roma<br>Roma<br>00133 |
| Italy | CESC della Provincia di Padova | Presso Azienda Ospedaliera di Padova_Via Giustiniani 1 Padova<br>Padova<br>35128 |
| Italy | Comitato Etico dell'Università Cattolica del Sacro Cuore e annesso Policlinico "A. Gemelli" | Largo Agostino Gemelli, 8 AOU Senese – Policlinico Agostino Gemelli<br>Roma<br>Roma 00168 |
| Italy | Comitato Etico della Provincia di Modena | Via del Pozzo, 71 Modena Modena<br>41124 |
| Italy | Comitato Etico Indipendente dell'Azienda Ospedaliero- Universitaria Policlinico S. Orsola-Malpighi BO | Via Pietro Albertoni, 15 Bologna<br>Bologna<br>40138 |
| Italy | Comitato Etico IRCCS Ospedale S. Raffaele di Milano | Via Olgettina, 60 Milano<br>Milano<br>20132 |

| | | |
|---|---|---|
| Italy | Comitato Etico Lazio 1 | Circonvalazione Gianicolense, 87<br>Roma<br>Roma<br>00152 |
| Italy | Comitato Etico Locale per la Sperimentazione Clinica dell'Azienda Ospedaliera Luigi Sacco di Milano | Via G. B. Grassi, 74 Padiglione 20-UO Farmacia<br>Milano Milano 20157 |
| Italy | Comitato Etico MIlano Area C | Piazza Ospedale Maggiore 3.. Milano<br>Milano<br>20162 |
| Italy | Comitato Etico per la Sperimentaz Clinica dei Medicinali A. O. U. Careggi di Firenze | Largo Brambilla, 3<br>Padiglione 15 Piastra dei Servizi Piano terra, stanza 31<br>Firenze<br>Firenze<br>50134 |
| Italy | Comitato Etico Referente per l'area di Pavia | Viale Camillo Golgi 19 Pavia<br>Pavia<br>27100 |
| Italy | Comitato Etico Regionale della Liguria | Largo Rosanna Benzi 10 Farmacia Ospedaliera Genova<br>16132 |
| Italy | Comitato Etico Scientifico dell'Azienda Ospedaliera Universitaria Policlinico Gaetano Martino | Via Consolare Valeria Messina<br>Messina<br>98125 |
| Italy | Istituto Clinico Humanitas | Via Manzoni, 56 Rozzano<br>Milano<br>20089 |
| Korea, Republic of | IRB of Asan Medical Center | 88 Olympic-ro 43-gil, Songpa-gu Seoul<br>Gyeonggi-do<br>05505 |
| Korea, Republic of | IRB of CHA Bundang Medical Center, CHA University | 59, Yatap-ro, Bundang-gu, Seongnam-si, Gyeonggi-do<br>13496 |
| Korea, Republic of | IRB of Dong-A University Medical Center | 26, Daesingongwon-ro, Seo-gu IRB, Dong A university Hospital Busan<br>Not Applicable<br>49201 |

| | | |
|---|---|---|
| Korea, Republic of | IRB of Korea University Ansan Hospital | 123, Jeokgeum-ro Danwon-gu Ansan-si Gyeonggi-do 15355 |
| Korea, Republic of | IRB of Kyungpook National University Hospital | 20F, JinSeck tower<br>Kyungpook National University Hospital kidney internal medicine clinical trial department<br>115, Dongdeok-ro, Jung-gu Daegu<br>Not Applicable<br>41944 |
| Korea, Republic of | IRB of Pusan National University Hospital | 179 Gudeok-ro, Seo-gu Busan<br>Not Applicable<br>49241 |
| Korea, Republic of | IRB of Samsung Medical Center | 81, Irwon-ro, Gangnam-gu Study coordinator room, B3, Annex Building, Samsung Medical Center<br>Seoul<br>Not Applicable 06351 |
| Korea, Republic of | IRB of Seoul National University Bundang Hospital | 172, Dolma-ro, Bundang-gu IRB, 3th floor, Healthcare Innovation Park Seongnam-si Gyeonggi-do 13605 |
| Korea, Republic of | IRB of Seoul National University Hospital | 101 Daehak-ro, Jongno-gu Seoul<br>Not Applicable<br>03080 |
| Korea, Republic of | IRB of Severance Hospital, Yonsei University Health System | No. 31 Office, Pediatric Oncology Clinic, Yonsei Cancer Hospital,<br>50-1 Yonsei-ro, Seodaemun-gu, Seoul<br>Not Applicable 03722 |
| Korea, Republic of | IRB of The Catholic University of Korea, St. Vincent's Hospital | 93, Jungbu-daero, Paldal-gu Suwon-si Gyeonggi-do<br>16247 |
| Mexico | CEI Hospital La Mision | Av. del Hospital 112 1° y 2° piso<br>Col. Sertoma Monterrey<br>Nuevo León 64718 |
| Mexico | Comite Bioetico para la Investigacion Clinica (CBIC) | Puebla No. 422, Despacho 4, Col. Roma Sur<br>Mexico<br>Distrito Federal<br>6700 |

| | | |
|---|---|---|
| Mexico | Comite de Etica de la Fac de Med de la UANL y Hospital Universitario "Dr. Jose Eleuterio Gonzalez" | Av. Francisco I. Madero Pte. s/n y Dr. E. Aguirre Pequeño, Col. Mitras Centro<br>Monterrey<br>Nuevo León 64460 |
| Mexico | Comite de Etica en Investigacion | Av Modesto Arreola #917 Ote. Col Centro<br>Monterrey<br>Nuevo León<br>64000 |
| Mexico | Comité de Ética en Investigación de Accelerium, S. de R.L. de C.V. | Modesto Arreola 917 Monterrey<br>Nuevo Leon<br>64000 |
| Mexico | Comité de Ética en Investigación de la Clínica Borda, S.A. de C.V. | Avenida Álvaro Obregón No. 107<br>Col. Centro<br>Cuernavaca<br>Morelos 62000 |
| Mexico | Comité de Ética en Investigacon de Resultados Médicos, Desarrollo e Investigación, SC | Blvd. Everardo Márquez No. 200, 3er Piso. Col Periodistas Pachuca<br>Hidalgo<br>42060 |
| Spain | CEIC de Aragón (CEICA) | Avda. Gómez Laguna, 25 planta 3<br>Zaragoza<br>Zaragoza<br>50009 |
| Spain | CEIC Fundación Hospital Alcorcón | C/ Budapest,1 Servicio psiquiatria<br>Alcorcón Madrid 28922 |
| Spain | CEIC Hospital de Fuenlabrada | Unidad de Investigación, Planta 1a<br>Camino del Molino, 2 Fuenlabrada<br>Madrid 28942 |
| Spain | Hospital Clinic de Barcelona | Villarroel 170<br>Servicio de Farmacia<br>Sotano 6B, Planta Sotano, 3a Puerta<br>Barcelona<br>Barcelona<br>08036 |

| | | |
|---|---|---|
| Spain | Hospital Universitari de Girona Dr Josep Trueta | Avda. de Franca, s/n<br>9a planta Zona A<br>Unitat de Neuroimmunologia i Esclerosi Multiple (JNIEM) Girona<br>Girona<br>17007 |
| Spain | Hospital Universitario de La Princesa | Diego de León, 62<br>Fundación para la Investigación Biomédica<br>Madrid<br>Madrid<br>28006 |
| Spain | Hospital Universitario La Paz | Paseo de la Castellana, 261 Neurologia<br>Planta 11<br>Madrid<br>Madrid 28046 |
| Switzerland | Ethikkommission des Kantons St.Gallen (EKSG) | Flurhof 7 St.Gallen 9007 |
| Switzerland | Kantonale Ethikkommission Bern (KEK-Bern) | Murtenstrasse 31 Postfach 56<br>Hörsaaltrakt Pathologie, Eingang 43A, Büro H372 Bern<br>3010 |
| Switzerland | Kantonale Ethikkommission Zürich (KEK-Zürich) | Stampfenbachstrasse 121 Zürich<br>8090 |
| United Kingdom | Barts Health NHS Trust | Joint Research and Develpment Office<br>Queen Mary Innovation Centre 5 Walden Street<br>London Greater London E1 2EF |
| United States | Biomedical Research Alliance of New York, LLC (BRANY) | 1981 Marcus Ave, Suite 210 Lake Success<br>New York<br>11041 |
| United States | CGIRB - IBC Services | 1019 39th Avenue SE Suite 120<br>Puyallup Washington 98374 |
| United States | Copernicus Group IRB | 5000 CentreGreen Way Suite 200<br>Cary<br>North Carolina<br>27513 |

| | | |
|---|---|---|
| United States | Henry Ford Health Systems IRB | Attn: Investigational Drug Services<br>2799 W. Grand Blvd A-Basement<br>Pharmacy Detroit<br>Michigan 48202 |
| United States | Mayo Clinic IRB - Rochester | 200 First Street SW, CH 1-285, 201<br>Building, Room 4-60<br>PET Imaging<br>Rochester Minnesota 55905 |
| United States | McGuire Institutional Review Board | 1201 Broad Rock Blvd. GI/Hepatology<br>111N Richmond<br>Virginia<br>23249 |
| United States | Medical College of Wisconsin, Inc. | 9200 Wisconsin Avenue, FEC- 4675<br>Milwaukee<br>Wisconsin<br>53226 |
| United States | MUSC - IBC | 19 Hagood Avenue Charleston<br>South Carolina 29425 |
| United States | Northwestern University IRB | 675 N Saint Clair St Chicago<br>Illinois<br>60611 |
| United States | NYU School of Medicine IRB | 550 First Ave<br>1201 Smilow Building New York<br>New York<br>10016 |
| United States | Partners Human Research Committee | 116 Huntington venue Suite 1002<br>Boston Massachusetts 02116 |
| United States | Thomas Jefferson University | 1015 Chestnut Street Suite 1100<br>Philadelphia Pennsylvania<br>19107 |
| United States | University of California IRB | 3333 California Street Suite 315<br>University of California San Francisco<br>California<br>94118 |
| United States | University of California San Diego | 8950 Villa La Jolla Drive Suite A208<br>La Jolla<br>California<br>92037 |

| | | |
|---|---|---|
| United States | University of Chicago IRB | 5841 S. Maryland Avenue, I- 625, MC7132<br>Office of Clinical Research Section of Regulatory Compliance<br>Chicago Illinois 60637 |
| United States | University of Louisville IRB | Med Center One, Suite 200 University of Louisville<br>501 E. Broadway<br>Louisville<br>Kentucky 40202 |
| United States | University of Minnesota Institutional Review Board | 420 Delaware Street SE<br>D528 Mayo Memorial Building<br>Minneapolis<br>Minnesota<br>55455 |
| United States | University of North Carolina at Chapel Hill IRB | CB 7097 Medical School Bldg 52 105 Mason Farm Rd<br>Chapel Hill<br>North Carolina<br>27599 |
| United States | University of Utah IRB | 75 South 2000 East Room 211<br>Salt Lake City<br>Utah<br>84112 |
| United States | University of Wisconsin- Madison, Health Sciences Institutional Board | 800 University Bay Drive Suite 105<br>University Bay Office Building Madison<br>Wisconsin 53792 |
| United States | UVA Institutional Review Board for Health Sciences Research | One Morton Drive Suite 400, Box 5 P.O. Box 800483 Charlottesville Virginia 22908 |
| United States | Vanderbilt University Medical Center IRB | 1313 21st Avenue South 504 Oxford House Nashville<br>Tennessee<br>37232 |
| United States | Washington University in St. Louis IRB | 660 South Euclid Avenue Campus Box 8223<br>St. Louis Missouri 63110 |

| | | |
|---|---|---|
| United States | Weill Cornell Medical College IRB | 1300 York Avenue New York<br>New York<br>10021 |
| United States | WIRB | 1019 39th Avenue SE Suite 120<br>Puyallup Washington<br>98374 |

***Supplemental Table 4: List of independent ethics committees / institutional review boards (Laurel)***

| Country | Authority/Committee Name | Authority/Committee Address |
|---|---|---|
| Brazil | CEP da Faculdade de Medicina do ABC/SP | Avenida Lauro Gomes, 2000 Vila Sacadura Cabral<br>Santo André<br>Sao Paulo<br>09060-870 |
| Brazil | CEP da Faculdade Una de Uberlandia | Alameda Paulina Malgonari, 59 Uberlandia<br>Minas Gerais<br>38411-206 |
| Brazil | CEP da Fundação Hospital Amaral Carvalho - SP | Rua Doutor Miranda Júnior, 16 Vila Assis<br>Jaú<br>Sao Paulo<br>17210-300 |
| Brazil | CEP da Universidade da Região de Joinville - UNIVILLE | Rua Paulo Malschitzki, 10 Joinville<br>Santa Catarina 89219-710 |
| Brazil | CEP da Universidade Federal de São Paulo / UNIFESP / EPM | Rua Botucatu, 572 - Conjunto 14 - 1o andar<br>Vila Clementino<br>São Paulo<br>Sao Paulo 04023-061 |
| Brazil | CEP do Hospital de Clinicas da Universidade Federal do Parana - HCUFPR / PR | Rua General Carneiro, 181 Alto da Glória<br>Curitiba<br>Paraná<br>80060-900 |
| Brazil | CEP do Hospital Moinhos de Vento/ RS | Rua Ramiro Barcelos, 910<br>Sala 902 - Centro Clínico Ramiro - Moinhos de Vento<br>Porto Alegre<br>Rio Grande do Sul<br>90035-001 |
| Brazil | CEP do Hospital Sírio Libanês | Rua Dona Adma Jafet, 91 São Paulo<br>Sao Paulo<br>01308-050 |
| Brazil | CEP do Hospital Universitário Gaffrée e Guinle/HUGG/UNIRIO | Rua Mariz e Barros, 775 - 2o andar<br>Tijuca<br>Rio de Janeiro<br>Rio do Janeiro 20270-004 |

| | | |
|---|---|---|
| Brazil | CEP do Hospital Universitário<br>Professor Edgard Santos - UFBA | Rua Augusto Viana, s/n Canela<br>Salvador<br>Bahia<br>40110-060 |
| Brazil | CEP do Hospital Universitario<br>Walter Cantidio - HUWC / CE | Rua Capitao Francisco Pedro, 1290 - Sala 12<br>Rodolfo Teofilo<br>Fortaleza<br>Ceará 60430-370 |
| Brazil | CEP do Instituto de Assistência<br>Médica ao Servidor Público<br>Estadual - IAMSPE | Rua Pedro de Toledo, 1800 - 3o andar - Ala Central<br>Vila Clementino<br>São Paulo<br>Sao Paulo 04039-901 |
| Brazil | CEP Investiga - Instituto de<br>Pesquisas | Avenida Romeu Tortima, 739 - Cidade Universitária<br>Campinas<br>Sao Paulo<br>13084-791 |
| Canada | Capital Health Authority Research<br>Ethics Boards | 5790 University Avenue Room 240<br>Halifax<br>Nova Scotia<br>B3H 1V7 |
| Canada | Hamilton Health Sciences<br>Integrated Review Board | 1280 Main Street West HSC 4N11<br>Hamilton<br>Ontario<br>L8S 4L8 |
| Canada | Mount Sinai Hospital | 600 University Ave Infectious Control Research,<br>Room 171<br>Toronto<br>ON<br>M5G 1X5 |
| Canada | University of Western Ontario | The University of Western Ontario<br>Office of Research<br>1151 Richmond Street London<br>Ontario N6A 3K7 |
| Canada | WIRB | 1019 39th Avenue SE Suite 120<br>Puyallup Washington<br>98374 |
| Czech<br>Republic | Eticka komise Fakultni nemocnice<br>Hradec Kralove | Sokolska 581 Hradec Kralove 50005 |

| | | |
|---|---|---|
| Czech Republic | Eticka komise Fakultni nemocnice u sv. Anny v Brne | Pekarska 53 Brno<br>656 91 |
| Czech Republic | Eticka komise FN a LF UP Olomouc | I.P.Pavlova 6 Olomouc 775 20 |
| Czech Republic | Eticka komise Nemocnice Na Bulovce | Budinova 67/2<br>Nemocnice Na Bulovce Gynekologicko-porodnicka klinika<br>Praha 8-Liben<br>180 81 |
| Czech Republic | Multicentricka eticka komise IKEM a TN | Videnska 800 Praha<br>140 59 |
| Hungary | Egeszsegugyi Tudomanyos Tanacs Klinikai Farmakologiai Etikai Bizottsaga | Alkotmany u.25 Budapest<br>1054 |
| India | Drug Trial Ethics Committee, Dayanand Medical College & Hospital | Department of Pediatrics Christian MEdical College and Hosital<br>Brown Road<br>Ludhiana Punjab 141008 |
| India | Ethics Committee | Ethics Committee, Dispur Hospital<br>Ganeshguri<br>Guwahati<br>Assam 781006 |
| India | Ethics Committee of the KLE University | Nehru Nagar JNMC Campus Belgaum Karnataka<br>590010 |
| India | Ethics Committee, M. S. Ramaiah Medical College and Hospitals | MSR Nagar, MSRIT Post<br>New BEL Road<br>M S Ramaiah Memorial Hospital Bangalore<br>Karnataka<br>560054 |
| India | Ethics Committee, Osmania Medical College | Koti<br>Hyderabad Andhra Pradesh 500095 |
| India | Institutional Ethics Committee | Maulana Azad Medical college,Bahadur Shah Zafar Marg<br>3 rd floor,Room no 306B New delhi<br>Delhi 110002 |
| India | Institutional Ethics committee I | Acharya Donde Marg, Parel,<br>Mumbai Maharashtra |

| | | |
|---|---|---|
| | | 400012 |
| India | Institutional Ethics Committee,<br>Deccan College of Medical Science | Deccan College of Medical Science,<br>Owaisi Hospital & Research Center,<br>Kanchanbagh Hyderabad Andhra Pradesh 500058 |
| India | Institutional Ethics committee,<br>Midas Multispeciality Hospital Pvt<br>Ltd. | Midas Height<br>07 Central Bazar road Ramdaspeth,<br>Nagpur<br>Maharashtra<br>440010 |
| India | Institutional Ethics Committee,<br>Poona Medical Research<br>Foundation pune | E4-C to E4-F, 4th Floor, Fifth Avenue<br>Pune<br>Maharashtra<br>411001 |
| India | Manipal University Ethics<br>Committee | 7th floor, Kasturba Hospital, Attavar<br>Mangalore<br>Karnataka<br>575001 |
| India | Nirmal Hospital Private Limited<br>Ethics Committee | Nirmal Hospital Private Limited Ethics Committee<br>Ring Road<br>Surat<br>Gujarat 395002 |
| India | P.D. Hinduja National Hospital &<br>Medical Research Centre | Veer Savarkar Marg Mahim Mumbai<br>Maharashtra<br>400 016 |
| India | Pushpawati Singhania Research<br>Institute for Liver, Renal &<br>Digestive Diseases | Press Enclave Marg Sheikh Sarai - II New Delhi<br>Delhi<br>110017 |
| India | S. R. Kalla Memorial Ethical<br>Committee for Human Research | 78, Dhuleshwar Garden, Behind HDFC Bank<br>Sardar Patel Marg, C Scheme Jaipur<br>Rajasthan 302001 |
| India | Shree Giriraj Hospital Research<br>Ethics Committee | 150 Feet Ring Rd, 27 Navjyot Park Main Road<br>Amin marg Cross Road Rajkot<br>Gujarat 360005 |
| India | Sparsh Hospital Ethics Committee | Sparsh Hospital & Critical Care (P) Ltd<br>A/407, Sahid Nagar Bhubaneswar<br>Orissa 751007 |

| | | |
|---|---|---|
| Israel | Assaf Harofeh MC Ethics Committee | Zerifin<br>Assaf Harofeh Center Rishon Lezion<br>75141 |
| Israel | Bnai Zion MC Ethics Committee | 47 golomb St. POB 4940 Haifa<br>31048 |
| Israel | Hadassah MC Ethics Committee | Kiryat Hadassah POB 12000 Jerusalem 91120 |
| Israel | Holy Family MC Ethics Committee | POB 8 HaGallil st Nazareth 16100 |
| Israel | Rambam Health Care Campus Ethics Committee | 8 Haaliyya St., Bat Galim POB 9602<br>Haifa<br>3109601 |
| Israel | Shaare Zedek MC Ethics Committee | 12 Hans Bayt Bait Vagan POB 3235 Jerusalem<br>9103102 |
| Israel | Wolfson MC Ethics Committee | 62 Halochamim St. POB 5<br>Holon<br>58100 |
| Italy | Comitato Etico Lazio 2 | Via Primo Carnera, 1 Roma<br>Roma<br>00142 |
| Italy | Comitato Etico MILANO Area C | Piazza Ospedale Maggiore n.3, Building 7, Floor 5<br>Milano<br>Milano<br>20162 |
| Italy | Comitato Etico Palermo 1 | Via del Vespro 127 Palermo<br>Palermo<br>90129 |
| Italy | Comitato Etico Provinciale della Provincia di Brescia | P.le Spedali Civili, 1 Brescia<br>Brescia<br>25123 |
| Mexico | Comite de Etica de la Fac de Med de la UANL y Hospital Universitario "Dr. Jose Eleuterio Gonzalez" | Av. Francisco I. Madero Pte. s/n y Dr. E. Aguirre Pequeño, Col. Mitras Centro<br>Monterrey<br>Nuevo León<br>64460 |
| Mexico | Comité de Ética del Centro de Investigación Clínica del Pacífico, S.A. de C.V. | La Nao #4, Cons. 1206, Colonia Magallanes Acapulco<br>Guerrero<br>C.P. 39670 |

| | | |
|---|---|---|
| Mexico | Comité de Ética en Investigación de Accelerium, S. de R.L. de C.V. | Modesto Arreola 917 Col. Centro Monterrey<br>Nuevo Leon<br>64000 |
| Mexico | Comité de Ética en Investigación de Hospital Misión | Avenida del Hospital No 112 Avenida del Hospital No. 112, 1° y 2° piso<br>Col. Sertoma<br>Monterrey<br>Nuevo León<br>64718 |
| Mexico | Comité de Ética en Investigación de la Clínica Borda, S.A. de C.V. | Avenida Álvaro Obregón No. 107<br>Col. Centro<br>Cuernavaca<br>Morelos 62000 |
| Mexico | Comité de Ética en Investigación de México Centre for Clinical Research S.A. de C.V. | Amores No. 709 Col. Del Valle, México, Distrito Federal, C.P. 03100. México<br>Mexico<br>Distrito Federal 03100 |
| Mexico | Instituto Nacional de Ciencias Medicas y Nutricion Salvador Zubiran | Vasco de Quiroga 15 Col. Belisario Domínguez Sección XVI, Tlalpan Mexico<br>Distrito Federal<br>14080 |
| Slovakia | Eticka komisia Bratislavskeho samospravneho kraja | Sabinovska 16 Bratislava<br>820 05 |
| Slovakia | Eticka komisia Kosickeho samospravneho kraja | Namestie Maratonu mieru 1 Kosice<br>04266 |
| Slovakia | Eticka komisia Presovskeho samospravneho kraja | Namestie mieru 2 Presov<br>08001 |
| Slovakia | Eticka komisia pri Fakultnej nemocnici Nitra | Spitalska 1 Nitra 95001 |
| Slovakia | Eticka komisia Univerzitnej nemocnice Bratislava | Ruzinovska 6 Bratislava 82606 |
| South Africa | Pharma Ethics | 123 Amcor Road Lyttelton Manor Centurion Pretoria<br>Gauteng<br>0157 |
| Ukraine | LEC Central City Clinical Hospital | 114, Hetmana Mazepy Str. Ivano-Frankivsk<br>76005 |

| | | |
|---|---|---|
| Ukraine | LEC CI of Kyiv RC Regional Clinical Hospital #2 | Nesterivskiy Lane 13/19 Kyiv 04053 |
| Ukraine | LEC CNE Lviv Regional Clinical Hospital | 7, Chernihivska St. Lviv 79010 |
| Ukraine | LEC Communal Institution Odesa Regional Clinical Hospital | 26, Ac. Zabolotnyi St. Odesa 65025 |
| Ukraine | LEC LLC Treatment and Diagnostic Center ADONIS Plus | 8-B, Raisy Okipnoyi St. LLC Treatment-Diagnostic Center ADONIS plus Kyiv 02002 |
| Ukraine | LEC Main Military Clin.Hospital | 18, Gospital'na Str. Kyiv 01133 |
| Ukraine | LEC Poltava Reg.Clin.Hosp.n.a.M.V.Sklifosovsko go | 23, Shevchenka St., Poltava 36011 |
| Ukraine | LEC SI Divisional Clinical Hospital of Uzhgorod Station of ST&BA LZ | 71, Mynaiska St. Uzhgorod 88009 |
| Ukraine | LEC Sumy Regional Council Sumy Reg Clin Hospital | 28, Troitska Str. Sumy 40022 |
| Ukraine | LEC Vinnytsia M.I.Pyrogov Regional Clinical Hospital | 46, Pyrogova St. Vinnytsia 21018 |
| Ukraine | LEC Zakarpatian Regional Clin.Hosp. n.a.A.Novak | 22 Peremohy St. Uzhgorod 88018 |
| United States | Baylor College of Medicine | One Baylor Plaza Mail Stop: BCM315 Houston Texas 77030 |
| United States | CGIRB - IBC Services | 1019 39th Avenue SE Suite 120 Puyallup Washington 98374 |
| United States | Copernicus Group IRB | 5000 CentreGreen Way Suite 200 Cary North Carolina 27513 |
| United States | Cornell University Institutional Review Board for Human Participants | 395 Pine Tree Road, Suite 320 Cornell University Ithaca New York 14850 |

| | | |
|---|---|---|
| United States | Henry Ford Health Systems IRB | Attn: Investigational Drug Services<br>2799 W. Grand Blvd A-Basement Pharmacy Detroit<br>Michigan 48202 |
| United States | Mayo Clinic IRB - Rochester | 200 First Street SW, CH 1-285, 201 Building, Room 4-60<br>PET Imaging<br>Rochester<br>Minnesota 55905 |
| United States | McGuire Institutional Review Board | 1201 Broad Rock Blvd. GI/Hepatology 111N<br>Richmond<br>Virginia<br>23249 |
| United States | Medical College of Wisconsin, Inc. | 9200 Wisconsin Avenue, FEC- 4675<br>Milwaukee<br>Wisconsin<br>53226 |
| United States | Northwestern University IRB | 675 N Saint Clair St Chicago<br>Illinois<br>60611 |
| United States | Thomas Jefferson University | 1015 Chestnut Street Suite 1100 Philadelphia<br>Pennsylvania<br>19107 |
| United States | University of California - Irvine IRB | 5171 California Avenue Suite 150<br>Irvine<br>California<br>92697 |
| United States | University of California IRB | 3333 California Street Suite 315<br>University of California San Francisco California<br>94118 |
| United States | University of Minnesota Institutional Review Board | 420 Delaware Street SE<br>D528 Mayo Memorial Building Minneapolis<br>Minnesota<br>55455 |
| United States | University of North Carolina at Chapel Hill IRB | CB 7097 Medical School Bldg 52 105 Mason Farm Rd<br>Chapel Hill<br>North Carolina<br>27599 |

| | | |
|---|---|---|
| United States | University of Texas Southwestern Investigational Review Board | 5323 Harry Hines Blvd.... Dallas<br>Texas<br>75390 |
| United States | University of Utah IRB | 75 South 2000 East Room 211<br>Salt Lake City<br>Utah<br>84112 |
| United States | UVA Institutional Review Board for Health Sciences Research | One Morton Drive Suite 400, Box 5 P.O. Box 800483<br>Charlottesville Virginia<br>22908 |
| United States | VA Long Beach IRB | 5901 E. Seventh Street Long Beach<br>California<br>90822 |
| United States | WIRB | 1019 39th Avenue SE Suite 120<br>Puyallup Washington<br>98374 |

***Supplemental Table 5: List of independent ethics committees / institutional review boards (Gardenia)***

| Country | Authority/Committee Name | Authority/Committee Address |
|---|---|---|
| Belgium | CHU de Liège | Domaine Universitaire du Sart Tilman<br>Batiment B 35<br>Liege<br>B-4000 |
| Belgium | Comite d ethique UCL St Luc Brussel | Avenue Hippocrate 55.14 Tour Harvey<br>niveau 0 Bruxelles<br>1200 |
| Belgium | Comité Local d'Ethique Hospitalier - C.H.U.<br>St.-Pierre | Bâtiment Direction Rue Haute 322<br>Bruxelles<br>1000 |
| Belgium | Ethisch Comité Universitair Ziekenhuis Gent | De Pintelaan 185<br>UZ Gent - Ethisch comite 1K4 Ghent<br>B-9000 |
| Belgium | Ethische Commissie - Algemeen Ziekenhuis<br>Sint- Elisabeth Herentals | Nederrij 133 Herentals 2200 |
| Belgium | GZA Ziekenhuizen | Oosterveldlaan 24 Wilrijk<br>2610 |
| Belgium | Imeldaziekenhuis | Imeldalaan 9 Bonheiden 2820 |
| Belgium | Universitair Ziekenhuis Brussel Commissie<br>Medische Ethiek | Laarbeeklaan 101 Brussels<br>1090 |
| Canada | Comité d'éthique de la recherche | 143 Wolfe Street Levis<br>Quebec<br>G6V 3Z1 |
| Canada | Conjoint Health Research Ethics Board | Room 9, Heritage Medical Research Buildig<br>Faculty of Medicine University of Calgary<br>Calgary<br>Alberta T2N 4N1 |
| Canada | CSSS Champlain Charles-Le Moyne REB | 3120, boul. Taschereau local AN-0132<br>Greenfield-Park Quebec<br>J4V 2H1 |
| Canada | Hamilton Health Sciences Integrated<br>Review Board | 1280 Main Street West HSC 4N11<br>Hamilton<br>Ontario |

| | | |
|---|---|---|
| | | L8S 4L8 |
| Canada | Health Research Ethics Board of Alberta | 1500-10104 103 Ave NW Edmonton<br>Alberta<br>T5J 4A7 |
| Canada | Humber River Hospital Research Ethics Board | 1235 Wilson Avenue Office of Research<br>Toronto<br>Ontario<br>M3M 0B2 |
| Canada | Maisonneuve-Rosemont hospital REB | 5415 l'Assomption Blvd Montreal<br>Quebec<br>H1T 2M4 |
| Canada | University of Alberta Health Research Ethics Board | 308 Campus Tower 8625 – 112 St.<br>Edmonton<br>Alberta<br>T6G 1K8 |
| Canada | University of Saskatchewan Biomedical REB | Box 5000 RPO University Saskatoon<br>Saskatchewan<br>S7N 4J8 |
| Canada | University of Western Ontario | The University of Western Ontario<br>Office of Research<br>1151 Richmond Street London<br>Ontario N6A 3K7 |
| Canada | Western University Office of Research Ethics Board | 1391 Western Road London<br>Ontario<br>N6G 1G9 |
| Canada | WIRB | 1019 39th Avenue SE Suite 120<br>Puyallup Washington<br>98374 |
| Czech Republic | Eticka komise - Krajska zdravotni a.s. | Socialni pece 3316/12A Usti nad Labem<br>40113 |
| Czech Republic | Eticka komise Fakultni nemocnice Hradec Kralove | Sokolska 581 Hradec Kralove 50005 |
| Czech Republic | Eticka komise IKEM a FTNsP | Videnska 800 Praha 4 - Krc 140 59 |
| Czech Republic | Eticka komise ISCARE I.V.F a.s. | Jankovcova 1569/2c Praha 7<br>17004 |
| Czech Republic | Eticka komise Krajska nemocnice T. Bati a.s. | Havlickovo nabrezi 600 Zlin<br>76275 |

| | | |
|---|---|---|
| Czech Republic | Eticka komise Mestska nemocnice Ostrava | Nemocnicni 898/20a Ostrava<br>70200 |
| Czech Republic | Eticka komise Nemocnice ve Frydku-Mistku p.o. | El. Krasnohorske 321 Frydek-Mistek 73801 |
| Czech Republic | Eticka komise Oblastni nemocnice Kladno a.s. | Vancurova 1548 Kladno<br>27259 |
| Czech Republic | Eticka komise Pardubicke krajske nemocnice | Kyjevska 44 Pardubice 53301 |
| Czech Republic | Eticka komise pro multicentricke klinicke hodnoceni Fakultni nemocnice v Motole | V Uvalu 84 Praha 5 15000 |
| Czech Republic | Eticka komise Vojenske nemocnice Brno | Zabrdovicka 3 Brno<br>636 00 |
| Hungary | Egeszsegugyi Tudomanyos Tanacs Klinikai Farmakologiai Etikai Bizottsaga | Alkotmany u.25 Budapest<br>1054 |
| Hungary | Ethik-Kommission der Medizinischen Fakultaet der Ernst-Moritz-Arndt-Universitaet Greifswald | Felix-Hausdorff-Str. 3 Greifswald<br>Mecklenburg Vorpommern 17475 |
| Israel | Barzilai MC Ethics Committee | 2 Hahistadrut St. Ashkelon<br>78278 |
| Israel | Chaim Sheba MC Ethics Committee | Tel Hashomer Ramat Gan 5265601 |
| Israel | Kaplan MC Ethics Committee | Pasternak way St. POB 1<br>Rehovot<br>76100 |
| Israel | Rabin MC Ethics Committee | 39 Jabotinsky St Ground floor Beilinson Campus Raphael Recanati Genetic Inst. Blg<br>Petach Tikva<br>4941492 |
| Israel | Sapir MC, Meir Hospital Ethics committee | 59 Tshernichovski St Kfar-Saba<br>4428126 |
| Israel | Tel Aviv Sourasky | 6 Weizman St. Tel Aviv 6423906 |
| Israel | The Baruch Padeh MC, Poriya Ethics Committee | The Baruch Padeh Medical Center<br>Poriya Cardiovascular Institute Mobile Post, Lower Galilee Tiberias<br>15208 |

| | | |
|---|---|---|
| Italy | Azienda Ospedaliera Universitaria Policlinico Tor Vergata | Viale Oxford, 81<br>Azienda Ospedaliera Univ Pol Tor Vergata - Dermatologia<br>Roma<br>Roma<br>00133 |
| Italy | Comitato Etico Area Vasta Nord Ovest | Via Roma 67 Pisa<br>Pisa<br>56126 |
| Italy | Comitato Etico Aziendale dell'Azienda Ospedaliero- Universitaria S. Maria della Misericordia di Udine | Via Colugna, 50 Udine<br>Udine<br>33100 |
| Italy | Comitato Etico CESC per la sperimentazione clinica delle province di Treviso e Belluno | Borgo Cavalli 42 Treviso<br>Treviso<br>31100 |
| Italy | Comitato Etico dell'Università Cattolica del Sacro Cuore e annesso Policlinico "A. Gemelli" | Largo Agostino Gemelli, 8 AOU Senese – Policlinico Agostino Gemelli<br>Roma Roma 00168 |
| Italy | Comitato Etico della Provincia di Modena | Via del Pozzo, 71 Modena Modena<br>41124 |
| Italy | Comitato Etico di Area Vasta Centro | Largo Brambilla, 3<br>Padiglione 15 Piastra dei Servizi Piano terra, stanza 31<br>Firenze<br>Firenze<br>50134 |
| Italy | Comitato Etico Indipendente dell'Azienda Ospedaliero- Universitaria Policlinico S. Orsola-Malpighi BO | Via Pietro Albertoni, 15 Bologna<br>Bologna<br>40138 |
| Italy | Comitato Etico Interaziendale Città della Salute Torino | Corso Bramante 88-90 Torino<br>Torino<br>10126 |
| Italy | Comitato Etico Interaziendale Milano Area A | Via G. B. Grassi, 74 Milano<br>Milano<br>20157 |
| Italy | Comitato Etico IRCCS Ospedale S. Raffaele di Milano | Via Olgettina, 60 Milano<br>Milano<br>20132 |

| | | |
|---|---|---|
| Italy | Comitato Etico Lazio 1 | Circonvalazione Gianicolense, 87 Roma<br>Roma<br>00152 |
| Italy | Comitato Etico Milano Area 2 | Via Francesco Sforza n. 28 Milano<br>Milano<br>20122 |
| Italy | Comitato Etico MILANO Area C | Piazza Ospedale Maggiore n.3, Building 7, Floor 5<br>Milano<br>Milano<br>20162 |
| Italy | Comitato Etico Palermo 2 | Viale Strasburgo 233 Palermo<br>Palermo<br>90145 |
| Italy | Comitato Etico per la Sperimentazione Clinica della Provincia di Padova | Via Berchet 10 Padova Padova<br>35131 |
| Italy | Comitato Etico Referente per l'area di Pavia | Viale Camillo Golgi 19 Pavia<br>Pavia<br>27100 |
| Italy | Comitato Etico Regionale della Liguria | Largo Rosanna Benzi 10 Farmacia Ospedaliera Genova<br>Genova<br>16132 |
| Italy | Comitato Etico Scientifico dell'Azienda Ospedaliera Universitaria Policlinico Gaetano Martino | Via Consolare Valeria Messina<br>Messina<br>98125 |
| Italy | Comitato Etico Unico per la Provincia di Parma | Via Gramsci, 14 Parma<br>Parma<br>43126 |
| Italy | IRCCS Ospedale Casa Sollievo della Sofferenza | Viale Cappuccini<br>San Giovanni Rotondo Foggia<br>71013 |
| Italy | Istituto Clinico Humanitas | Via Manzoni, 56 Rozzano<br>Milano<br>20089 |
| Korea, Republic of | IRB of Ajou University Hospital | 164, Worldcup-ro Yeongtong-gu Suwon<br>Gyeonggi-do 16499 |

| | | |
|---|---|---|
| Korea,<br>Republic of | IRB of Asan Medical Center | 88 Olympic-ro 43-gil, Songpa-gu Seoul<br>Gyeonggi-do<br>05505 |
| Korea,<br>Republic of | IRB of CHA Bundang Medical Center, CHA University | 59, Yatap-ro, Bundang-gu, Seongnam-si,<br>Gyeonggi-do<br>13496 |
| Korea,<br>Republic of | IRB of Dong-A University Medical Center | 26, Daesingongwon-ro, Seo-gu IRB, Dong A university Hospital<br>Busan<br>Not Applicable 49201 |
| Korea,<br>Republic of | IRB of Hanyang University Guri Hospital | 153, Gyeongchun-ro Guri-si<br>Gyeonggi-do<br>11923 |
| Korea,<br>Republic of | IRB of Kangbuk Samsung Hospital | 29, Saemunan-ro, Jongno-gu Seoul<br>Not Applicable<br>03181 |
| Korea,<br>Republic of | IRB of Keimyung University Dongsan Hospital | 56, Dalseong-ro, Jung-gu clinical trial pharmacy,3F, ByeolGwan<br>Daegu<br>Not Applicable 41931 |
| Korea,<br>Republic of | IRB of Konyang University Hospital | 158, Gwanjeodong-ro, Seo-gu,<br>Daejeon<br>Not Applicable<br>35365 |
| Korea,<br>Republic of | IRB of Korea University Ansan Hospital | 123, Jeokgeum-ro Danwon-gu Ansan-si<br>Gyeonggi-do 15355 |
| Korea,<br>Republic of | IRB of Kyung Hee University Hospital | 23, Kyungheedae-ro, Dongdaemun-gu<br>Seoul<br>Not Applicable 02447 |
| Korea,<br>Republic of | IRB of Kyungpook National University Chilgok Hospital | 807, Hoguk-ro, Buk-gu Daegu<br>Gyeongsangbuk-do 41404 |
| Korea,<br>Republic of | IRB of Kyungpook National University Hospital | 20F, JinSeck tower<br>Kyungpook National University Hospital kidney internal medicine clinical trial department<br>115, Dongdeok-ro, Jung-gu Daegu<br>Not Applicable<br>41944 |

| | | |
|---|---|---|
| Korea, Republic of | IRB of Pusan National University Hospital | 179 Gudeok-ro, Seo-gu Busan<br>Not Applicable<br>49241 |
| Korea, Republic of | IRB of Samsung Medical Center | 81, Irwon-ro, Gangnam-gu<br>Study coordinator room, B3, Annex Building, Samsung Medical Center<br>Seoul<br>Not Applicable 06351 |
| Korea, Republic of | IRB of Seoul Metropolitan Government Seoul National University Boramae Medical Center | 209, 2F, Lotte Tower, 51, Boramae-ro 5-gil, Dongjak-gu Seoul<br>Not Applicable<br>156-849 |
| Korea, Republic of | IRB of Seoul National University Bundang Hospital | 172, Dolma-ro, Bundang-gu IRB, 3th floor, Healthcare Innovation Park Seongnam-si Gyeonggi-do 13605 |
| Korea, Republic of | IRB of Seoul National University Hospital | 101 Daehak-ro, Jongno-gu Seoul<br>Not Applicable<br>03080 |
| Korea, Republic of | IRB of Severance Hospital, Yonsei University Health System | No. 31 Office, Pediatric Oncology Clinic, Yonsei Cancer Hospital,<br>50-1 Yonsei-ro, Seodaemun-gu, Seoul<br>Not Applicable 03722 |
| Korea, Republic of | IRB of The Catholic University of Korea, St. Vincent's Hospital | 93, Jungbu-daero, Paldal-gu Suwon-si Gyeonggi-do<br>16247 |
| Korea, Republic of | IRB of Yeungnam University Hospital | 170 Hyeonchung-ro, Nam-gu Daegu<br>Not Applicable<br>42415 |
| Korea, Republic of | IRB of Yonsei University Wonju Severance Christian Hospital | 5th floor, Mun Chnag Mo memorial Building, 20 Ilsan-Ro Wonju-si Gangwon-do<br>26426 |
| Philippines | Cardinal Santos Medical Center - IRB | 21 Wilson St. Greenhills San Juan City<br>1500 |
| Singapore | National Healthcare Group Domain Specific Review Board | No. 6 Commonwealth Lane GMTI building Level 6 Singapore<br>149547 |

| | | |
|---|---|---|
| Singapore | SingHealth Centralised Institutional Review Board | Block A, 7 Hospital Drive SingHealth Research Facilities, #03-01<br>Singapore<br>169611 |
| South Africa | Pharma Ethics | 123 Amcor Road Lyttelton Manor Centurion Pretoria Gauteng<br>0157 |
| South Africa | University of Pretoria | Tswelopele Building opposite BMW building<br>Level 4, Room 4-59<br>Pretoria<br>Gauteng 0002 |
| South Africa | University of the Free State | Faculty of Health Sciences<br>Block D, Room 115, Francois Retief Building<br>Dean's Division, Nelson Mandela Drive<br>Bloemfontein Free State 9301 |
| Spain | CEIC Fundación Hospital Alcorcón | C/ Budapest,1 Servicio psiquiatria<br>Alcorcón Madrid 28922 |
| Spain | CEIC Hospital de Fuenlabrada | Unidad de Investigación, Planta 1a<br>Camino del Molino, 2 Fuenlabrada<br>Madrid 28942 |
| Spain | CEIC Hospital General Universitario de Alicante | c/ Pintor Baeza, 12<br>3a planta del edificio gris Alicante<br>Alicante<br>03010 |
| Spain | Fundación Biomédica Complexo Hospitalario Universitario de Vigo | C/ Meixoeiro, s/n Vigo<br>Pontevedra 36200 |
| Spain | Hospital Clinic de Barcelona | Villarroel 170<br>Servicio de Farmacia<br>Sotano 6B, Planta Sotano, 3a Puerta<br>Barcelona<br>Barcelona<br>08036 |

| | | |
|---|---|---|
| Spain | Hospital Universitari de Girona Dr Josep Trueta | Avda. de Franca, s/n<br>9a planta Zona A<br>Unitat de Neuroimmunologia i Esclerosi Multiple (JNIEM) Girona<br>Girona<br>17007 |
| Spain | Hospital Universitario Clinico San Carlos | Servicio Farmacología Clínica 1a planta, ala Norte, Puerta G c/ Doctor Martin Lagos, s/n Madrid<br>Madrid 28040 |
| Spain | Hospital Universitario de La Princesa | Diego de León, 62<br>Fundación para la Investigación Biomédica<br>Madrid<br>Madrid<br>28006 |
| Spain | Hospital Universitario La Paz | Paseo de la Castellana, 261 Neurologia<br>Planta 11<br>Madrid<br>Madrid 28046 |
| Switzerland | Ethikkommission des Kantons St.Gallen (EKSG) | Flurhof 7 St.Gallen 9007 |
| Switzerland | Kantonale Ethikkommission Bern (KEK-Bern) | Murtenstrasse 31<br>Postfach 56<br>Hörsaaltrakt Pathologie, Eingang 43A, Büro H372<br>Bern<br>3010 |
| Switzerland | Kantonale Ethikkommission Zürich (KEK-Zürich) | Stampfenbachstrasse 121 Zürich<br>8090 |
| United Kingdom | R&D - Royal Devon and Exeter NHS Foundation Trust | Barrack Road Wonford Exeter Devon<br>EX2 5DW |
| United Kingdom | R&D - The Newcastle upon Tyne Hospitals NHS Foundation Trust | Royal Victoria Infirmary Queen Victoria Road Newcastle Upon Tyne Newcastle North Humberside NE1 4LP |
| United Kingdom | R&D - University Hospital Southampton NHS Foundation Trust | Mailpoint 18, Southampton General Hospital<br>Tremona Road<br>Southampton Hampshire SO16 6YD |

| | | |
|---|---|---|
| United Kingdom | R&D Cambridge University Hosptials NHS Foundation Trust | Research and Development Department<br>Box 277, Addenbrooke's Hospital Hills Road<br>Cambridge<br>Cambridgeshire<br>CB2 0QQ |
| United Kingdom | R&D Kings Health Partners | Clinical Trials Office<br>Floor 16, Tower Wing, Guys Hospital<br>Great Maze Pond<br>London<br>Greater London<br>SE1 9RT |
| United Kingdom | R&D Royal Bournemouth and Christchurch Hospitals NHS Foundation Trust | Castle Lane East Bournemouth Dorset<br>BH7 7DW |
| United Kingdom | R&D The Leeds Teaching Hospitals NHS Trust | Leeds Teaching Hospitals NHS Trust<br>Research & Development<br>34 Hyde Terrace<br>Leeds<br>West Yorkshire LS2 9LN |
| United Kingdom | R&D UCLH NHS Foundation Trust | Joint Research Office<br>1st Floor Maple House Suite B 149<br>Tottenham Court Road London<br>Greater London<br>W1T 7DN |
| United Kingdom | R&D University Hospitals Coventry and Warwickshire NHS Trust | Walgrave General Hospital Clifford Bridge Road<br>Coventry<br>West Midlands CV2 2DX |
